# Association of CSF α-Synuclein Seed Amplification Assay Positivity with Disease Progression and Cognitive Decline: A Longitudinal Alzheimer’s Disease Neuroimaging Initiative Study

**DOI:** 10.1101/2024.07.16.24310496

**Authors:** Duygu Tosun, Zachary Hausle, Pamela Thropp, Luis Concha-Marambio, Jennifer Lamoureux, Russ Lebovitz, Leslie M. Shaw, Andrew B. Singleton, Michael W. Weiner, the Alzheimer’s Disease Neuroimaging Initiative, Cornelis Blauwendraat

## Abstract

**INTRODUCTION:** CSF α-synuclein seed amplification assay (SAA) is a sensitive and specific tool for detecting Lewy body (LB) co-pathology in AD.

**METHODS:** 1637 cross-sectional and 407 longitudinal CSF samples from ADNI were tested with SAA. We examined longitudinal dynamics of Aβ, α-synuclein seeds, and p-tau181, along with global and domain-specific cognition in stable SAA+, stable SAA−, and those who converted to SAA+ from SAA−.

**RESULTS:** SAA+ individuals had faster cognitive decline than SAA−, notably in MCI, and presented with earlier symptom onset. SAA+ conversion was associated with CSF Aβ42-positivity but did not impact progression of either Aβ42 or p-tau181 status. Aβ42, p-tau181, and α-syn SAA were all strong predictors of clinical progression, particularly Aβ42. In vitro α-syn SAA kinetic parameters were associated with participant demographics, clinical profiles, and cognitive decline.

**DISCUSSION:** These results highlight the interplay between Aβ and α-synuclein and their association with disease progression.

## INTRODUCTION

Alzheimer’s disease (AD) and Lewy body disease (LBD), characterized by the pathological deposition of amyloid-beta (Aβ) and alpha-synuclein (α-syn), respectively, are commonly identified at autopsy. Up to half (25-50%) of autopsy cases exhibit Lewy body (LB) co-pathology in sporadic early- and late-onset AD [1–4], familial/inherited AD [5], and Down’s Syndrome AD cases [6]. Pathological coexistence implies a potential interplay between Aβ and α-syn in the human brain. Despite the unique stereotypical progression of each pathology [7–9], evidence suggests that these pathways potentially may overlap at later disease stages [1–4], implicating a synergistic process known as ‘crosstalk’.

Crosstalk has been observed in neurodegenerative diseases and can occur by impaired cellular clearance processes, impaired protein homeostasis, synergy of disease related pathways, or when amyloidogenic proteins such as Aβ, tau, and α-syn interact and cause aggregation [10–13]. Each pathologic deposition occurs in distinct, observable locations in the brain: Aβ plaques are extracellular, tau neurofibrillary tangles (NFTs) are intracellular, and LB aggregates are in vesicles and exosomes [14,15]. Despite this, interaction of these proteins may intersect at later disease stages, potentially exacerbating disease progression. Our understanding of crosstalk in living organisms, particularly in humans, remains limited. As such, the mechanism, and dynamics of interaction between Aβ and α-syn, and the subsequent impact on disease progression are areas of active research. With the recently developed cerebrospinal fluid (CSF) α-syn seed amplification assay (SAA) technology, there is now in vivo evidence that individuals with both LB (α-syn) and AD (Aβ and tau) pathology exhibit faster cognitive decline than those with only LB or AD pathology [16–19].

In fact, evidence from autopsy studies showed that the Aβ and tau pathologies only account for a portion of the observed variance in cognitive decline [20] while co-pathologies lower the threshold for clinical symptoms of AD [21]. Accordingly, while current treatments with anti-Aβ antibodies have been shown to slow cognitive decline, their impact is relatively modest, reducing the rate of decline by approximately 25-40% [22–24]. This underscores the possibility that additional pathologies may play a critical role. Given that α-syn is the most commonly observed co-pathology in AD [25], the presence of α-syn pathology could help explain the variability in cognitive decline that is not accounted for by Aβ and tau alone.

Recently, analysis of cross-sectional CSF samples from the Alzheimer’s Disease Neuroimaging Initiative (ADNI) study using SAA demonstrated an association between the presence of misfolded α-syn and various factors such as age, disease stage, burden of AD pathology, and rates of longitudinal cognitive decline [19]. We recently expanded the ADNI CSF α-syn SAA analysis by incorporating longitudinal time points, aiming for a better understanding of longitudinal downstream effects resulting from Aβ and α-syn pathologies in ADNI participants. To our knowledge, this is the first study to incorporate longitudinal AD biomarker data with longitudinal SAA data in an AD cohort where extensive longitudinal follow-up allowed us to identify individuals who progressed from SAA-negative (SAA−) to SAA-positive (SAA+).

Here, in the context of AD co-pathologies, we hypothesize that SAA-positivity would correspond to greater rates of cognitive decline and earlier onset of cognitive impairment. Further, we postulate that the emergence of α-syn pathology is dependent on pre-existing AD Aβ pathology, and the apolipoprotein E (*APOE*) ε4 allele exerts a significant influence over this interplay, since ε4 has been increasingly recognized as a common genetic risk factor for both AD and LBD [26–29]. *APOE* contributes to progression and cognitive decline in Parkinson’s [30], and ε4 has been shown to worsen α-syn pathology in AD+LB brains [31], suggesting a role in modulating crosstalk. Lastly, the dichotomous outcome derived from the qualitative SAA approach poses a limitation. To address this, we investigate the utility of SAA kinetic parameters as quantitative indicators for the burden of α-syn seeds in CSF or the propagation of LB pathology. Thus, we assess the association of the quantitative SAA kinetic parameters with clinical characteristics, biomarker data, and cognitive outcome measures.

## METHODS

### Study design and participants

Data was obtained from the ADNI database (https://adni.loni.usc.edu/). The ADNI was launched in 2003 as a public-private partnership, led by Principal Investigator Michael W. Weiner, MD. The primary goal of ADNI has been to test whether serial magnetic resonance imaging (MRI), positron emission tomography (PET), other biological markers, and clinical and neuropsychological assessment can be combined to measure the progression of mild cognitive impairment (MCI) and early AD. For up-to-date information, see www.adni-info.org. This study employed a longitudinal examination of biomarker, demographic, clinical, and cognitive data.

The sample included all ADNI 1-3 cohort participants who had available CSF samples (N=1637). The participant pool consisted of cognitively unimpaired (CU) individuals, individuals with MCI, and individuals clinically diagnosed with dementia due to AD.

In summary, at the time of enrollment, participants in the ADNI study were aged between 55 and 90 years, had a study partner to provide an independent evaluation of functioning, and were proficient in English or Spanish. For CU individuals, enrollment criteria included a Mini–Mental State Examination (MMSE) score between 24 and 30, a Clinical Dementia Rating (CDR) of 0, absence of depression, MCI, and dementia.

MCI participants were required to have MMSE scores between 24 and 30, a subjective memory complaint, objective memory loss (adjusted for education) as measured by the Wechsler Memory Scale Logical Memory II, a CDR of 0.5, no significant impairment in other cognitive domains, essentially preserved daily living activities, and no dementia.

Participants diagnosed with dementia due to AD met the criteria with MMSE scores between 20 and 26, a CDR of 0.5 or 1.0, and fulfilled the NINCDS/ADRDA criteria for probable AD.

Exclusion criteria at the time of ADNI study enrollment included significant neurological disease apart from AD (including PD and DLB), contraindications to neuroimaging or other ADNI protocols, neuroimaging evidence of infection, infarction, lacunes, or other focal lesions, psychiatric disorders, including psychotic features, alcohol abuse, significant systemic illness or unstable medical condition, laboratory abnormalities that could interfere with the study, use of certain psychoactive medications, and participation in other clinical trials.

### CSF **α**-synuclein seed amplification assay processing

CSF samples were initially gathered into collection tubes provided to every participating ADNI site. These were then transferred to polypropylene tubes and frozen on dry ice within an hour of collection. The samples were subsequently shipped overnight, still on dry ice, to the ADNI Biomarker Core laboratory at the University of Pennsylvania Medical Center. Upon their arrival at the ADNI Biomarker Core laboratory, the CSF samples were thawed and aliquoted into 0.5 mL cryotubes for long-term storage at −80°C.

The CSF α-syn SAA testing was carried out by the Amprion Clinical Laboratory (CLIA ID No. 05D2209417; CAP No. 8168002) using a method clinically validated and compliant with Clinical Laboratory Improvement Amendment (CLIA) standards.

For the analysis, each pristine aliquot of CSF was tested in three technical replicates within a 96-well plate. The 100 µL reaction mixture was composed of 100 mM PIPES pH 6.5, 0.5 M NaCl, 0.1% sarkosyl, 10 µM ThT, 0.3 mg/mL recombinant α-syn, 40 µL CSF, and two silicon nitride beads [18]. Positive and negative quality control samples were included in each plate to ensure assay accuracy.

The plates were sealed with an optical adhesive film and placed into a BMG LABTECH FLUOstar Ω Microplate Reader. They were incubated at 42°C, with cycles of one minute of shaking followed by 14 minutes of rest. Fluorescence was recorded after each shake, using an excitation wavelength 440 nm and emission wavelength of 490 nm. After a total incubation period of 20 hours, the maximum fluorescence intensity for each well was recorded. An algorithm was then applied to the triplicate reading to categorize the result.

CSF samples were classified as follows: “PD/DLB-like Detected” if α-syn aggregates were identified with an aggregation profile consistent with Type 1 seeds observed in Parkinson’s Disease (PD) and Dementia with Lewy bodies (DLB); “MSA-like Detected” if α-syn aggregates matched Type 2 seeds typically seen in Multiple System Atrophy (MSA); or “Not Detected” if no α-syn aggregates were detected. Samples that did not yield a definitive result after two tests were classified as “Indeterminate”.

The processing of ADNI CSF α-syn SAA was done in two phases. Phase-1 data were processed in Q4 2023 and included 1,637 CSF samples from the latest CSF sample collection time point for each participant. Phase-2 data were processed in Q1 2024 and incorporated CSF samples from earlier collection time points specifically focusing on participants showing detectable seeding activity from Phase-1. These earlier time points were from 222 participants and included those classified as “PD/DLB-like Detected”, “MSA-like Detected”, “Indeterminate”, and those with postmortem neuropathological confirmation, totaling 407 additional CSF samples. CSF samples from earlier collection time points of participants whose most recent CSF samples were classified as “Not Detected” were not processed unless they were in the autopsy cohort. This decision assumed, supported by data from prior studies, that their earlier CSF samples would also likely be classified as ‘Not Detected’.

For the in vitro assay of SAA, the following five kinetic parameters (illustrated in Supplemental Figure S1) were estimated for each SAA+ replicate: 1) Time to Threshold (TTT, [hours]) – time in hours when the fluorescence signal reaches the lower patient classification threshold (1000 RFU); 2) Maximum Fluorescence (Fmax, [RFU]) – maximum of the reaction signal in relative fluorescence units (RFU); 3) AUC-Fluoro (RFU, [seconds]) – area under the signal versus time reaction curve in RFU; 4) Maximum Slope (Smax, [RFU, seconds]) – maximum of the derivative of the signal/time reaction curve in RFU/seconds; 5) Time to Smax (TSmax, [hours]) – the time in hours when the maximum slope occurs.

All CSF α-syn SAA analyses were performed with the analysts blinded to the participants’ demographic details, clinical profiles, and AD biomarker data. The integrity of the blinding was maintained by utilizing unique specimen identifiers for randomly assigned sample shipments.

### Assessments of CSF AD biomarkers

Pristine aliquots of CSF were examined using the Elecsys CSF Aβ42, CSF phospho-tau181 (p-tau181), and CSF total-tau electrochemiluminescence immunoassays (ECLIA) on a fully automated Elecsys cobas e 601 instrument, utilizing a single lot of reagents for each biomarker. The Roche Elecsys CSF immunoassays were used in accordance with a Roche Study Protocol at the ADNI Biomarker Laboratory, following the kit manufacturer’s instructions.

The analyses were carried out in a series of runs, with each sample run once (in singlicate) for each biomarker test, from November 17, 2016, through June 22, 2022. This process followed a standard new lot rollover protocol from the manufacturer, which involved repeated analyses of quality control samples.

The analyte measuring ranges from the lower technical limit to the upper technical limit for each biomarker were: 200 – 1700 pg/mL for the Elecsys CSF Aβ42 immunoassay, and 8 – 120 pg/mL for the Elecsys CSF p-tau181 immunoassay.

The AD CSF biomarker positivity was defined as “Aβ42+” if CSF Aβ42 was less than 980 pg/mL, and “p-tau181+” if CSF p-tau181 was greater than 24 pg/mL.

Subject ages at time of conversion to CSF Aβ42 positivity (i.e., Aβ age) were estimated using sampled iterative local approximation (SILA) on all ADNI subjects with CSF Aβ42 data available, as described in Betthauser et. al 2022 [32]. Records outside of the technical limits of the Elecsys CSF Aβ42 assay (lower technical limit of 200 pg/mL and upper technical limit of 1700 pg/mL) were excluded from the Aβ age estimation.

### Cognitive Assessments

The global cognitive assessments included the CDR - Sum of Boxes (CDR-SB), the Alzheimer’s Disease Assessment Scale - cognitive subscale 11-item (ADAS-Cog11), MMSE based on a 30-point questionnaire, and the Preclinical Alzheimer’s Composite Score (PACC). The domain-specific cognitive assessments included the composite measures of memory, executive function, and language [33]. Observations in domain-specific measures were excluded if the standard error of measurement for a given observation exceeded 0.6.

### Statistical Analysis

All statistical analyses and data preparation were conducted in R (version 4.4.0), except SILA Aβ age estimation, which was conducted in MATLAB. Holm–Bonferroni method, was used to correct for multiple comparisons, when applicable.

Subjects with any MSA-like samples were excluded from all analyses because of small sample size in this group, as detailed in the Results section. Indeterminate CSF α-syn SAA samples were discarded for all primary analyses. Samples were designated as SAA− (“Not Detected”) if no α-syn aggregates were detected, and SAA+ (“PD/DLB-like Detected”) if α-syn aggregates conformed to Type 1 seeds, typically observed in PD and DLB. Subjects who were SAA− after baseline were inferred to have been SAA− at all prior observations, as detailed in the Results section.

Subjects were classified as “converters” in each measure if they were negative in a measure and later were positive in that measure and remained positive at all subsequent observations. A converting subject’s conversion date was estimated as the midpoint between their last biomarker negative date and first biomarker positive date. Subjects who were positive in a measure and were then negative at any subsequent observations were classified as reverse converters and were excluded from analyses.

Subjects were classified as “Stable SAA−” if they had multiple CSF samples and were SAA− at their last observation, “Stable SAA+” if they were SAA+ at two or more observations and were not SAA− at any observation. Subjects that only had one CSF observation did not have enough information to sort into these groups and were excluded from analyses that involved these groupings.

Demographic, CSF biomarker, and cognitive measures were compared cross-sectionally for subjects across the Stable SAA−, SAA Converter, and Stable SAA+ groups listed, with observations before and after conversion included for SAA Converters. The most recent observation with SAA data available was used for both the Stable SAA− and Stable SAA+ groups.

Pairwise group differences were assessed. Binary variables for converters were compared before and after conversion using McNemar’s test, continuous variables using paired t-tests, and diagnosis using paired sign tests. For all other group comparisons, categorical comparisons between groups were performed using chi-squared tests when all categories had enough observations, and Fisher’s tests when one or more did not. All cognitive measures and continuous CSF Aβ42 were compared between groups with ANCOVAs adjusted for age, sex, years of education, diagnosis, and *APOE* ε4 genotype. Continuous CSF p-tau181 was compared between groups with ANCOVAs adjusted for age, sex, years of education, diagnosis, *APOE* ε4 genotype, and CSF Aβ42 status. Logistic regressions, adjusted for age, sex, years of education, and *APOE* ε4 genotype, were performed to compare group differences in CSF AD biomarker positivity.

Generalized Additive Mixed-effects Models (GAMMs) with penalized cubic regression spline were fit to assess the changes in longitudinal cognitive outcome measures, as a function of Aβ-time. Aβ-time at the cognitive assessment time was measured relative to the SILA-estimated Aβ-age at CSF Aβ42 positivity. GAMMs were fit separately for Stable SAA− and Stable SAA+ groups while accounting for differences in age, sex, years of education, and *APOE* ε4 genotype. Aβ-time at which Stable SAA− and Stable SAA+ groups reached a cognitive performance threshold defined as two standard deviations below the mean of CU participants were estimated via bootstrap resampling.

To evaluate the effects of transitioning to SAA positivity among SAA Converters, we first identified a reference group of Stable SAA− individuals. We matched their cognitive assessment time points with those of SAA Converters at their last SAA− evaluation. The matching criteria included age, sex, years of education, *APOE* ε4 genotype, and Aβ-time, using a 2-to-1 genetic matching approach. The duration of cognitive assessments prior to the last SAA− evaluation for SAA Converters was matched to the duration prior to the corresponding matched time point for the Stable SAA− reference group. Matching was stratified by clinical diagnostic groups (i.e., CU, MCI, and Dementia). We then applied piecewise mixed-effects regression models to the longitudinal cognitive data, setting a predefined breakpoint at t = 0. This breakpoint represents the estimated SAA conversion time for the SAA Converters and the midpoint between the matched time point and the subsequent assessment for the Stable SAA− reference group. The estimated cognitive decline rates before (t < 0) and after (t > 0) the breakpoint were compared between the SAA Converters and the Stable SAA− reference groups.

Survival analysis was conducted for conversion in four outcomes: SAA positivity, Aβ42 positivity, p-tau181 positivity, and clinical diagnosis progression (i.e., CU to MCI/Dementia or MCI to Dementia). Time for all outcomes was measured from baseline. Kaplan-Meier survival curves, stratified by the status of other outcomes at baseline and *APOE* ε4 genotype, were estimated for all outcomes. Cox proportional hazards models were fit for each outcome separately. Person-period coding was used to reflect change in outcome measures. CSF SAA status was included as time-variant predictors in models for CSF AD positivity and clinical diagnosis progression models. Similarly, CSF Aβ42 and p-tau181 statuses were included as time-variant predictors in models for CSF SAA positivity and clinical diagnosis progression models. Age at baseline, sex, and *APOE* £4 genotype were included in all models.

Next, we evaluated the extent to which the SAA kinetic parameters are associated with disease characteristics and risk factors, with a particular focus on the Stable SAA+ and SAA Converter cohorts. We assessed the independent effects of age, sex, *APOE* ε4 status, clinical diagnosis, and CSF Aβ42 positivity on SAA kinetic parameters, in a full model including all of these factors.

Association of SA kinetic parameters with CSF Aβ42 and p-tau181 levels were assessed using linear regression models adjusted for age, sex, *APOE* ε4 status, and clinical diagnosis, where p-tau181 model was further adjusted for CSF Aβ42 positivity. Similarly, the relationship between SAA kinetic parameters and cognitive outcome scores cross-sectionally at the first CSF time-point were assessed using linear regression models adjusted for age, sex, *APOE* ε4 status, and clinical diagnosis. To determine how changes in SAA kinetic parameters are associated with the progression of cognitive decline, we used linear mixed-effects models (LMM), with cognitive measure of interest as the outcome variable and time since the initial CSF sample collection, the SAA kinetic parameter, and their interaction as predictor variables. We adjusted these models for age, sex, educational years, and *APOE* ε4 status, incorporating random intercepts and slopes to account for correlations within subjects. We conducted these analyses for each SAA kinetic parameter and within each clinical diagnostic category independently. Finally, leveraging longitudinal kinetic data from the SAA Converter group, we repeated the association between changes in SAA kinetic parameters and cognitive decline rates, using time since the initial SAA+ measurement in the longitudinal models.

In all SAA kinetic parameter association analyses we included the total CSF protein concentration as a surrogate measurement for lipoproteins as it was shown to affect the kinetics of αsyn seed amplification in a concentration-dependent manner [34].

## RESULTS

### ADNI SAA study cohort characteristics

The initial set of CSF α-syn SAA analysis (Phase-1; Figure 1) comprised the latest CSF specimens from 1,637 participants who were part of the ADNI 1-3 studies. Out of these, 368 (22.5%) specimens exhibited PD/DLB-like α-syn seeding activity (i.e., SAA+), while three showed MSA-like α-syn seeding activity. No α-syn seeding aggregation was observed in 1,256 samples, which were thus classified as SAA−. The SAA outcomes were indeterminate for ten samples. These findings from the cross-sectional CSF α-syn SAA have been previously reported in detail [19].

**Figure 1:**
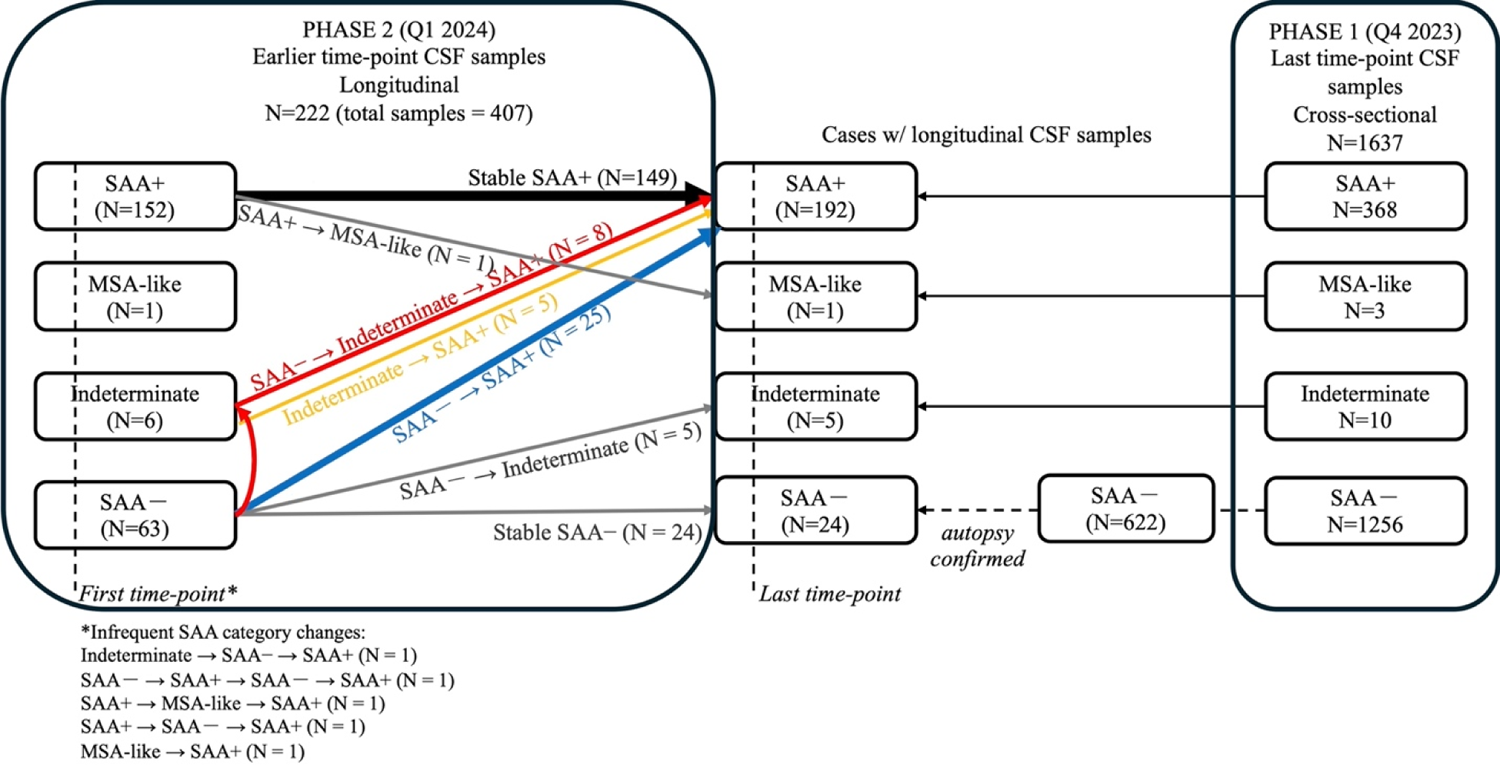
Graphical overview of the ADNI CSF α-syn Seed Amplification Assay (SAA) workflow. On the right, Phase-1 included SAA measurement of all ADNI 1-3 participants’ most recent time point, previously published (Tosun et al 2024). As follow-up in Phase-2 we included all previous CSF timepoints available from groups identified as 1) SAA+, 2) MSA-like, and 3) indeterminate in Phase-1 processing, and 4) autopsy cohort, totaling 222 participants with 407 samples.

The samples for the second set of longitudinal CSF α-syn SAA analysis (Phase-2; Figure 1) were selected from those with available CSF samples from previous study timepoints, totaling 819.

This selection was narrowed down to include only those identified as SAA+ (192 individuals), those with MSA-like α-syn seeding (one individual), or those with indeterminate seeding activity (five individuals) as determined by the initial cross-sectional analysis at their last CSF collection (Phase-1). Additionally, Phase-2 also included samples from all the participants from the ADNI autopsy sub-cohort, which led to inclusion of an additional 24 individuals who were identified SAA− at their last CSF collection. In total, Phase-2 comprised 407 longitudinal CSF samples from 222 distinct participants.

### Assessment of Longitudinal CSF α-syn SAA profiles

As depicted in Figure 1, the CSF samples from participants who were classified as SAA− at their final CSF collection (totaling 24 individuals) consistently tested SAA− at all preceding timepoints as well, suggesting stability of the SAA− findings retrospectively. Consequently, for those individuals who were determined to be SAA− at their most recent CSF collection according to Phase-1 analysis but were not selected for the longitudinal Phase-2 processing (N=598), their CSF samples from earlier time points were deemed as SAA−. Including the 24 SAA− samples from the longitudinal Phase-2, these individuals (N=622) were collectively classified as having a Stable SAA− status.

Among the 222 participants included in the longitudinal Phase 2 analysis, 63 were initially classified as SAA− at their first CSF time point. Of these, 24 remained consistently SAA−, thus described earlier as Stable SAA−. In contrast, 25 individuals progressed from SAA− to SAA+ by their final CSF time point. Additionally, there were eight participants who, while initially SAA−, had an interim CSF time point yielding indeterminate SAA results, but ultimately were found to be SAA+ by their final CSF sample. Another five began as SAA− and maintained this status up to their last CSF collection, which ended with an indeterminate SAA result. As one of the infrequent instances of fluctuating SAA categories, there was one participant who initially tested as SAA−, subsequently transitioned to SAA+, reverted to SAA−, and then returned to being SAA+ by the subsequent assessments.

Out of the six participants who initially presented with indeterminate SAA results, five were classified as SAA+ by the time of their last CSF collection. As one of the infrequent instances of fluctuating SAA categories, there was one participant who initially had an indeterminate SAA result, subsequently tested SAA−, and then transitioned to being SAA+ by the subsequent assessments. In all the analyses described below, CSF samples with indeterminate SAA results were excluded.

Among the 222 participants in the longitudinal Phase-2 study, 152 were initially classified as SAA+ at their first CSF collection. Of these, 149 consistently tested SAA+ across all study timepoints, i.e. stable SAA+. However, one participant changed from the SAA+ category to exhibiting MSA-like seeding patterns by their final CSF collection. The CSF specimen with MSA-like seeding was visibly discolored, likely due to blood contamination. Two rare cases of fluctuating SAA categories included one individual transitioning from SAA+ to MSA-like and then back to SAA+, and another who moved from SAA+ to SAA−, and then reverted to SAA+.

Additionally, there was a single case of a participant who initially presented with an MSA-like seeding pattern and subsequently tested SAA+ in their final CSF sample. The CSF specimen with MSA-like seeding was visibly discolored, likely due to blood contamination. For the analyses that follow, any individual who showed MSA-like α-syn seed aggregation at any time point, regardless of their initial or final CSF SAA status, was excluded. Moreover, the two individuals demonstrating the patterns SAA− → SAA+ → SAA− → SAA+ and SAA+ → SAA−→ SAA+ were also excluded from the study analyses. In total 5 out of 222 (2%) of the participants in the longitudinal Phase-2 study were excluded from the study analysis.

In total, 34 individuals presented with SAA− → SAA+ pattern. Converters averaged 2.5 years between their last visit with a SAA− result and their first visit with a SAA+ result, with a minimum of 0.9 years and a maximum of 6.1 years between those visits. The mid-point between their last visit with a SAA− result and their first visit with a SAA+ result was considered as the SAA conversion time. Among participants with a SAA− sample at baseline and follow up CSF samples, proportion of SAA Converters within baseline diagnosis groups of CU, MCI, and Dementia were 3.7% (11 out of 297), 6.7% (20 out of 299), and 5.1% (3 out of 59), respectively. Among SAA Converters, 1 out of 11 (9%) subjects who were CU at baseline progressed to MCI at their first visit after SAA conversion, and 10 out of 20 (50%) subjects who were MCI at baseline were diagnosed with dementia at their first visit after SAA conversion. One converter (5%) who was MCI at baseline was diagnosed as CU at their first visit after SAA conversion.

There were no significant differences in CSF levels of total protein, white blood cell count, and red blood cell count between groups with CSF samples categorized as SAA+, SAA−, and indeterminate.

### Cohort characteristics of Stable SAA−, SAA Converters, and Stable SAA+

Next, we assessed the demographic, biomarker, and clinical characteristics of SAA groups, including Stable SAA− group (N=622), SAA Converter group (N=34), and Stable SAA+ group (N=149), as shown in Table 1.

**Table 1:** Demographic, clinical, and biomarker characteristics of the individuals with CSF _-syn SAA− stable over time (Stable SAA−), individuals with CSF _-syn SAA+ stable over time (Stable SAA+), and individuals progressed from CSF _-syn SAA− to CSF _-syn SAA+ (SAA Converters). For SAA Converters, cohort characteristics are provided before and after conversion time points (i.e., last time point with CSF _-syn SAA− and first time point with CSF _-syn SAA+, respectively). n (%) are provided for dichotomized and categorical variables, median (IQR) for continuous variables. Missing data counts and percentages for clinical and biomarker data are provided in Supplemental Table S1.

| Characteristic | i. Stable SAA–<br>N = 622 | ii. SAA Converters<br>(Before), N = 34 | iii. SAA Converters<br>(After), N = 34 | iv. Stable SAA+<br>N = 149 | p<br>i vs. ii | p<sup>7</sup><br>ii vs. iii | p<br>iii vs. iv | p<br>i vs. iv |
| --- | --- | --- | --- | --- | --- | --- | --- | --- |
| <sup>1</sup>Diagnosis |  |  |  |  | 0.40 | 0.81 | 0.59 | <0.0 |
| CU | 269 (43%) | 11 (32%) | 11 (32%) | 36 (24%) |  |  |  |  |
| MCI | 213 (34%) | 15 (44%) | 10 (29%) | 52 (35%) |  |  |  |  |
| Dementia | 140 (23%) | 8 (24%) | 13 (38%) | 61 (41%) |  |  |  |  |
| <sup>2</sup>Age | 76 (71, 81) | 76 (66, 79) | 78 (70, 83) | 78 (73, 81) | 0.08 | <0.001 | 0.67 | 0.1 |
| <sup>1</sup>Male | 316 (51%) | 21 (62%) | 21 (62%) | 87 (58%) | 0.29 | N/A | 0.89 | 0.1 |
| <sup>3</sup>Ethnicity |  |  |  |  | 0.71 | N/A | N/A | 0.0 |
| Hispanic or Latino | 18 (2.9%) | 0 (0%) | 0 (0%) | 0 (0%) |  |  |  |  |
| Not Hispanic or Latino | 599 (96%) | 34 (100%) | 34 (100%) | 149 (100%) |  |  |  |  |
| Unknown | 5 (0.8%) | 0 (0%) | 0 (0%) | 0 (0%) |  |  |  |  |
| <sup>3</sup>Race |  |  |  |  | 0.13 | N/A | 0.01 | 0.0 |
| Asian | 4 (0.6%) | 0 (0%) | 0 (0%) | 4 (2.7%) |  |  |  |  |
| Black or African American | 24 (3.9%) | 1 (2.9%) | 1 (2.9%) | 4 (2.7%) |  |  |  |  |
| More than one race | 13 (2.1%) | 3 (8.8%) | 3 (8.8%) | 0 (0%) |  |  |  |  |
| White | 581 (93%) | 30 (88%) | 30 (88%) | 141 (95%) |  |  |  |  |
| <sup>2</sup>Years of education | 16.00 (14.00, 18.00) | 18.00 (16.00, 20.00) | 18.00 (16.00, 20.00) | 16.00 (14.00, 18.00) | 0.03 | N/A | 0.02 | 0.3 |
| <sup>3</sup>APOE ε4 genotype | | | | | 0.03 | N/A | 0.41 | 0.1 |
| Non-Carrier | 370 (59%) | 15 (44%) | 15 (44%) | 80 (54%) |  |  |  |  |
| Heterozygotes | 202 (33%) | 12 (35%) | 12 (35%) | 50 (34%) |  |  |  |  |
| Homozygotes | 50 (8.1%) | 7 (21%) | 7 (21%) | 19 (13%) |  |  |  |  |
| <sup>4</sup>CSF Aβ42 | 849 (591, 1,387) | 732 (580, 1,024) | 638 (516, 1,098) | 690 (495, 1,211) | 0.14 | 0.21 | 0.65 | 0.0 |
| <sup>6</sup>CSF Aβ42+ | 338 (57%) | 25 (74%) | 23 (70%) | 97 (66%) | 0.13 | >0.9 | 0.41 | 0.5 |
| <sup>5</sup>CSF p-tau181 | 24 (17, 35) | 27 (21, 38) | 27 (21, 43) | 24 (18, 34) | 0.04 | >0.9 | 0.01 | 0.5 |
| <sup>6</sup>CSF p-tau181+ | 287 (49%) | 20 (59%) | 22 (67%) | 72 (50%) | 0.41 | 0.37 | 0.10 | 0.1 |
| <sup>4</sup>ADAS-Cog11 | 13 (7, 22) | 15 (10, 23) | 21 (10, 29) | 19 (11, 30) | 0.16 | <0.001 | 0.90 | <0.0 |
| <sup>4</sup>CDR-SB | 1.0 (0.0, 3.0) | 1.0 (0.5, 3.4) | 2.0 (0.5, 4.9) | 2.0 (0.5, 5.0) | 0.37 | <0.01 | 0.36 | <0.0 |
| <sup>4</sup>PACC | -3 (-10, 1) | -6 (-10, -1) | -10 (-12, -1) | -8 (-15, -2) | <0.01 | <0.01 | 0.50 | <0.0 |
| <sup>4</sup>MMSE | 28.0 (25.0, 30.0) | 28.0 (25.0, 29.0) | 26.0 (23.0, 29.0) | 26.0 (22.0, 29.0) | 0.10 | <0.01 | 0.88 | <0.0 |
| <sup>4</sup>Executive function | 0.47 (0.01, 0.94) | 0.36 (-0.23, 0.65) | 0.26 (-0.06, 0.62) | 0.10 (-0.55, 0.64) | 0.05 | 0.22 | 0.21 | <0.0 |
| <sup>4</sup>Memory | 0.45 (-0.35, 1.10) | 0.13 (-0.35, 0.75) | -0.40 (-0.65, 0.64) | -0.11 (-0.83, 0.64) | 0.06 | <0.001 | 0.77 | <0.0 |
| <sup>4</sup>Language | 0.51 (0.08, 0.82) | 0.54 (-0.09, 0.81) | 0.44 (-0.17, 0.76) | 0.16 (-0.30, 0.62) | >0.9 | 0.04 | 0.25 | <0.0 |

<sup>1</sup> Pearson's Chi-squared test
<sup>2</sup> One-way ANOVA
<sup>3</sup> Fisher's exact test
<sup>4</sup> ANCOVA adjusted for age, sex, education, diagnosis, and APOE
<sup>5</sup> ANCOVA adjusted for age, sex, education, APOE, diagnosis, and CSF Aβ 42 status
<sup>6</sup> Logistic regression adjusted for age, sex, education, diagnosis, and APOE
<sup>7</sup> Paired t-test: all continuous variables; McNemar's test: all binary variables; paired sign test: diagnosis.

The Stable SAA+ group, when compared to Stable SAA− individuals, had a higher proportion of cognitively impaired individuals, lower levels of CSF Aβ42, and poorer scores in all global and domain-specific cognitive measures assessed in this study. Stable SAA+ group did not include any participants with Hispanic/Latino ethnic background and a greater frequence of Asian racial background but a lower frequence of Black/African American racial background in comparison to the Stable SAA−. Although these racial and ethnic differences between Stable SAA+ and Stable SAA− groups were significant (p=0.04), these results should be interpreted with caution given the low ethnoracial diversity in ADNI.

SAA Converters exhibited a higher level of educational attainment when compared to individuals who remained consistently Stable SAA− or Stable SAA+ throughout the ADNI study. A greater frequency of *APOE* ε4 homozygotes was observed among SAA Converters in comparison to Stable SAA− group. SAA Converters at their last visit with a SAA− result had increased levels of CSF p-tau181 and poorer performance on the PACC and executive function composite score relative to the Stable SAA− group. In addition, SAA Converters at their first visit with an SAA+ result presented with greater levels of CSF p-tau181 compared to the Stable SAA+ group. Furthermore, SAA Converters were more likely to be from non-white racial groups than those who were consistently Stable SAA+, though this result should be interpreted with caution due to the small sample size.

Lastly, between their last SAA− assessment and the subsequent SAA+ result, five SAA Converters who were previously diagnosed with MCI advanced to a clinical diagnosis of dementia due to AD. Three individuals from the MCI SAA Converter group developed dementia following their SAA+ conversion, while one individual initially classified as MCI SAA Converter later reverted to CU, as detailed in Supplemental Table S2. Relative to their cognitive performance at their last SAA− time point, SAA Converters demonstrated greater impairment in all global and domain-specific cognitive measures except the executive function composite measures at their initial SAA+ time point.

### Association of SAA-positivity with the rates of cognitive decline

After accounting for differences in age, sex, years of education, *APOE* ε4 genotype, and CSF Aβ 42 and p-tau181 levels, Stable SAA+ CU participants, compared to their Stable SAA− counterparts, experienced faster increases in ADAS-Cog11 and steeper decline in PACC, memory and executive function composite scores (Table 2). Similarly, Stable SAA+ MCI participants showed more rapid declines across all global and domain specific cognitive measures assessed, relative to Stable SAA− MCI participants, with a medium Cohen’s f^2^ effect size of 0.15 – 0.21 [35]. Only SAA-positivity association rapid cognitive decline in ADAS-Cog11, PACC, memory, executive function, and language measures within MCI survived Holm– Bonferroni correction.

**Table 2:**
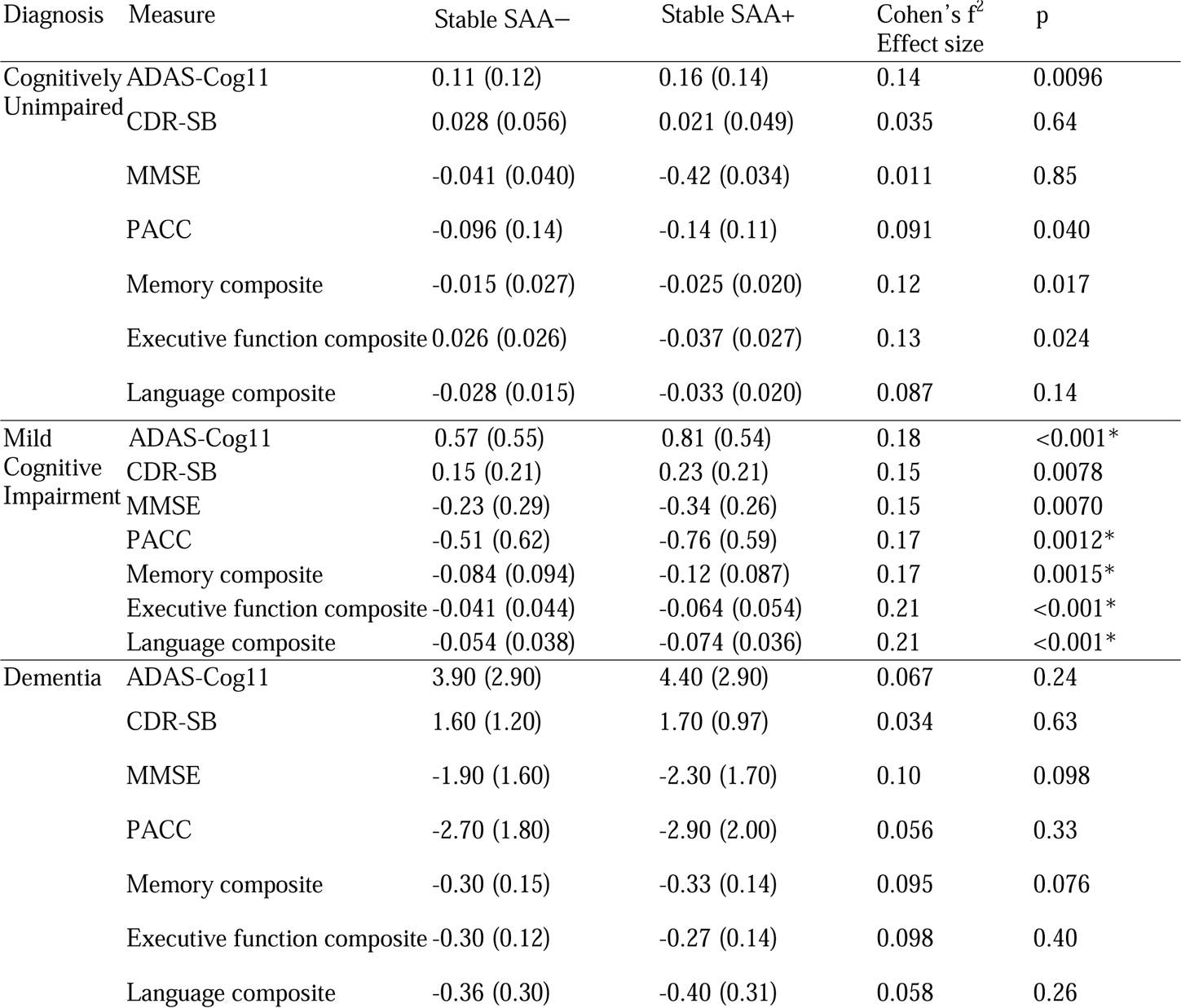
Rates of change in cognitive outcome measures for Stable SAA− and Stable SAA+ groups. Rates of change in each cognitive outcome measure were separately modeled for all groups. Estimated rates were compared across groups after adjusting for age, sex, gender, *APOE* genotype, and CSF Aβ42 and p-tau181 levels. Rates are listed as mean (SD). *Significance survived Holm–Bonferroni correction.

| Diagnosis | Measure | Stable SAA– | Stable SAA+ | Cohen's $f^2$<br>Effect size | p |
| --- | --- | --- | --- | --- | --- |
| Cognitively Unimpaired | ADAS-Cog11 | 0.11 (0.12) | 0.16 (0.14) | 0.14 | 0.0096 |
|  | CDR-SB | 0.028 (0.056) | 0.021 (0.049) | 0.035 | 0.64 |
|  | MMSE | -0.041 (0.040) | -0.42 (0.034) | 0.011 | 0.85 |
|  | PACC | -0.096 (0.14) | -0.14 (0.11) | 0.091 | 0.040 |
|  | Memory composite | -0.015 (0.027) | -0.025 (0.020) | 0.12 | 0.017 |
|  | Executive function composite | 0.026 (0.026) | -0.037 (0.027) | 0.13 | 0.024 |
|  | Language composite | -0.028 (0.015) | -0.033 (0.020) | 0.087 | 0.14 |
| Mild Cognitive Impairment | ADAS-Cog11 | 0.57 (0.55) | 0.81 (0.54) | 0.18 | <0.001* |
|  | CDR-SB | 0.15 (0.21) | 0.23 (0.21) | 0.15 | 0.0078 |
|  | MMSE | -0.23 (0.29) | -0.34 (0.26) | 0.15 | 0.0070 |
|  | PACC | -0.51 (0.62) | -0.76 (0.59) | 0.17 | 0.0012* |
|  | Memory composite | -0.084 (0.094) | -0.12 (0.087) | 0.17 | 0.0015* |
|  | Executive function composite | -0.041 (0.044) | -0.064 (0.054) | 0.21 | <0.001* |
|  | Language composite | -0.054 (0.038) | -0.074 (0.036) | 0.21 | <0.001* |
| Dementia | ADAS-Cog11 | 3.90 (2.90) | 4.40 (2.90) | 0.067 | 0.24 |
|  | CDR-SB | 1.60 (1.20) | 1.70 (0.97) | 0.034 | 0.63 |
|  | MMSE | -1.90 (1.60) | -2.30 (1.70) | 0.10 | 0.098 |
|  | PACC | -2.70 (1.80) | -2.90 (2.00) | 0.056 | 0.33 |
|  | Memory composite | -0.30 (0.15) | -0.33 (0.14) | 0.095 | 0.076 |
|  | Executive function composite | -0.30 (0.12) | -0.27 (0.14) | 0.098 | 0.40 |
|  | Language composite | -0.36 (0.30) | -0.40 (0.31) | 0.058 | 0.26 |

We next modeled the longitudinal trajectories of cognitive outcome measures as a function of Aβ-time, while adjusting for age, sex, years of education, and *APOE* ε4 genotype (Figure 2). We estimated the relative Aβ-time for the SAA+ and SAA− groups to reach a cognitive performance threshold defined as two standard deviations below the mean of CU participants. For the SAA− group, the time from CSF Aβ42 positivity (Aβ-time) to reach the cognitive performance threshold was as follows: 5.72 ± 0.47 years for ADAS-Cog11, 0.61 ± 1.06 years for MMSE, 1.77 ± 0.55 years for the PACC, 12.02 ± 0.39 years for memory function, 13.82 ± 0.44 years for executive function, and 16.41 ± 0.46 years for language function. In contrast, the SAA+ individuals reached the same cognitive performance thresholds 5.7 to 9.5 years earlier than their SAA− counterparts. The Aβ-time to threshold for the SAA+ group and significance of the differences compared to SAA− were as follows: −0.40 ± 1.68 years for ADAS-Cog11 (p=0.00049), −5.63 ± 1.11 years for MMSE (p<0.0001), −7.75 ± 1.05 years for PACC (p<0.0001), 4.00 ± 1.27 years for memory function (p<0.0001), 7.99 ± 1.37 years for executive function (p<0.0001), and 10.74 ± 1.75 years for language function(p=0.0028).

**Figure 2:**
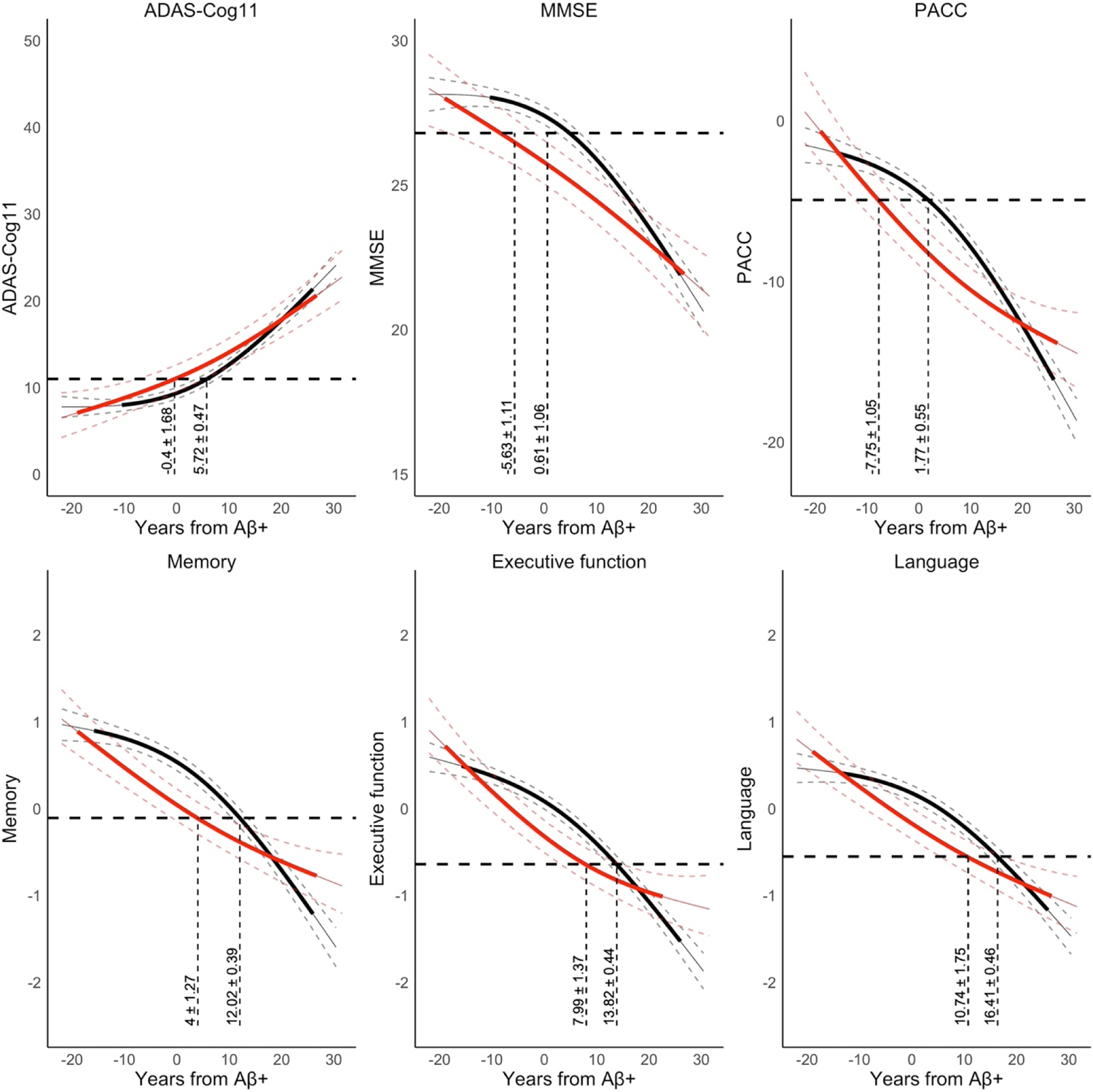
The longitudinal trajectories of cognitive outcome measures as a function of Aβ-time, while adjusting for age, sex, years of education, and *APOE* ε4 genotype. Aβ-time at the cognitive assessment time was measured relative to the SILA-estimated Aβ-age at CSF Aβ42 positivity. Horizontal dashed lines indicate the cognitive performance threshold defined as two standard deviations below the mean of CU participants.

Next, we compared SAA Converters (N=34) with a matched group of Stable SAA− individuals (Reference group) in a 2:1 ratio. Our aim was to assess their cognitive decline rates both before and after the critical conversion point, denoted as t=0 (Figure 3). For SAA Converters, t=0 represents the approximate time of conversion to SAA+, while for the Reference group, it aligns with the point where they were matched to the SAA Converters based on age, sex, years of education, *APOE* ε4 status, Aβ-time, and clinical diagnosis. After Holm–Bonferroni correction, MCI SAA Converters compared to the MCI Reference group showed a significantly accelerated decline in PACC (z=4.10 p<0.0001) and memory function (z=4.52 p<0.0001) after their estimated time of SAA conversion.

**Figure 3:**
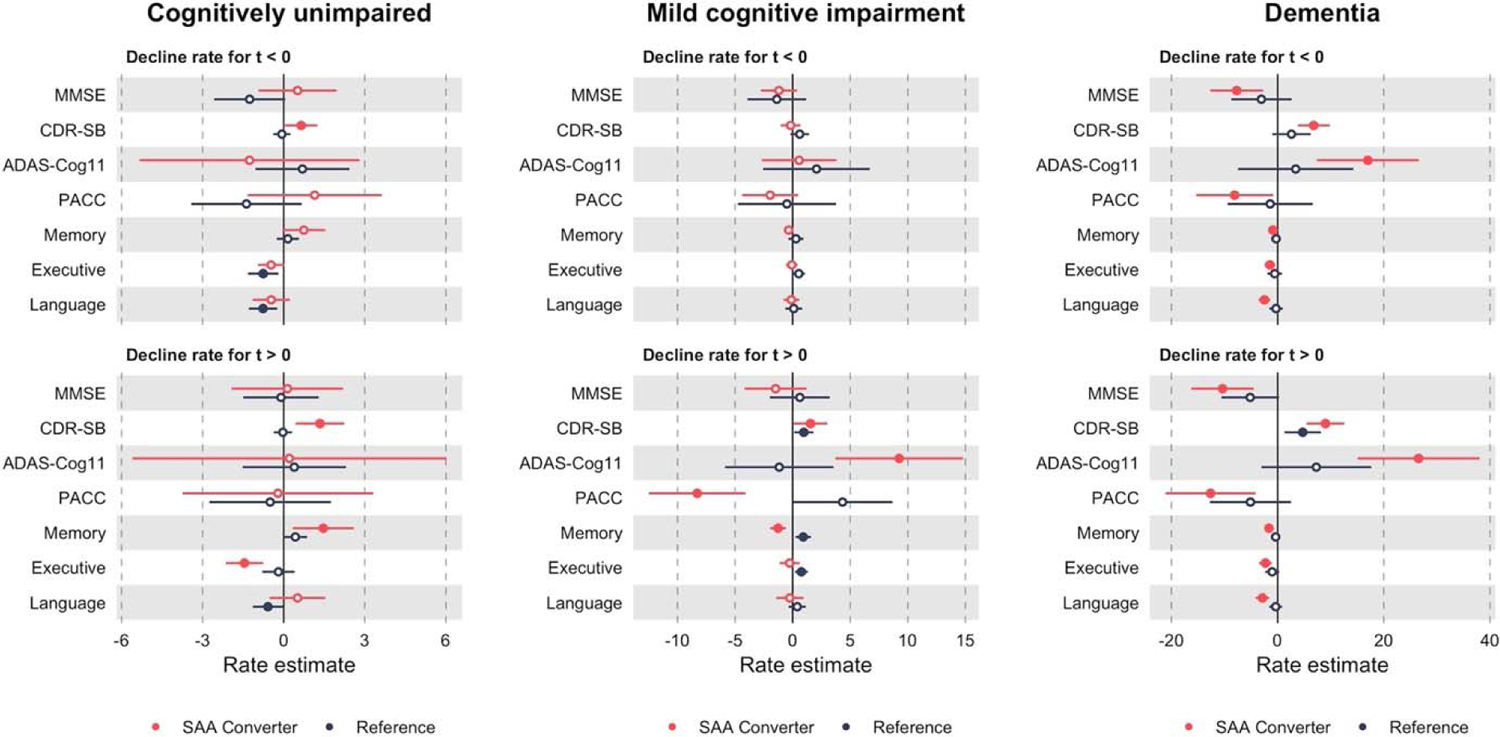
Cognitive decline rates for SAA Converters (N=34) and a matched group of Stable SAA− individuals (Reference group). For SAA Converters, t=0 represents the approximate time of conversion to SAA+, while for the Reference group, it aligns with the point where they were matched to the SAA Converters based on age, sex, years of education, *APOE* ε4 status, Aβ-time, and clinical diagnosis in a 2:1 ratio. Closed and open circles indicate p ≤ 0 and p > 0, respectively, for the estimated cognitive decline rates.

### SAA Conversion time relative to CSF A**β** time

The timing of the CSF Aβ42 pathology relative to SAA conversion is illustrated in Figure 4. Among the 34 SAA Converters, 70% (N=24; 7 out of 8 Dementia, 13 out of 15 MCI, and 4 out of 11 CU) were CSF Aβ42 positive before their SAA conversion time point. On average, Aβ-time (i.e., time from CSF Aβ42 positivity) for Dementia and MCI SAA Converters was 14.9 years and 9.4 years prior to their SAA conversion time, respectively, while Aβ-time for CU SAA Converters was 2.7 years after their SAA conversion time.

**Figure 4:**
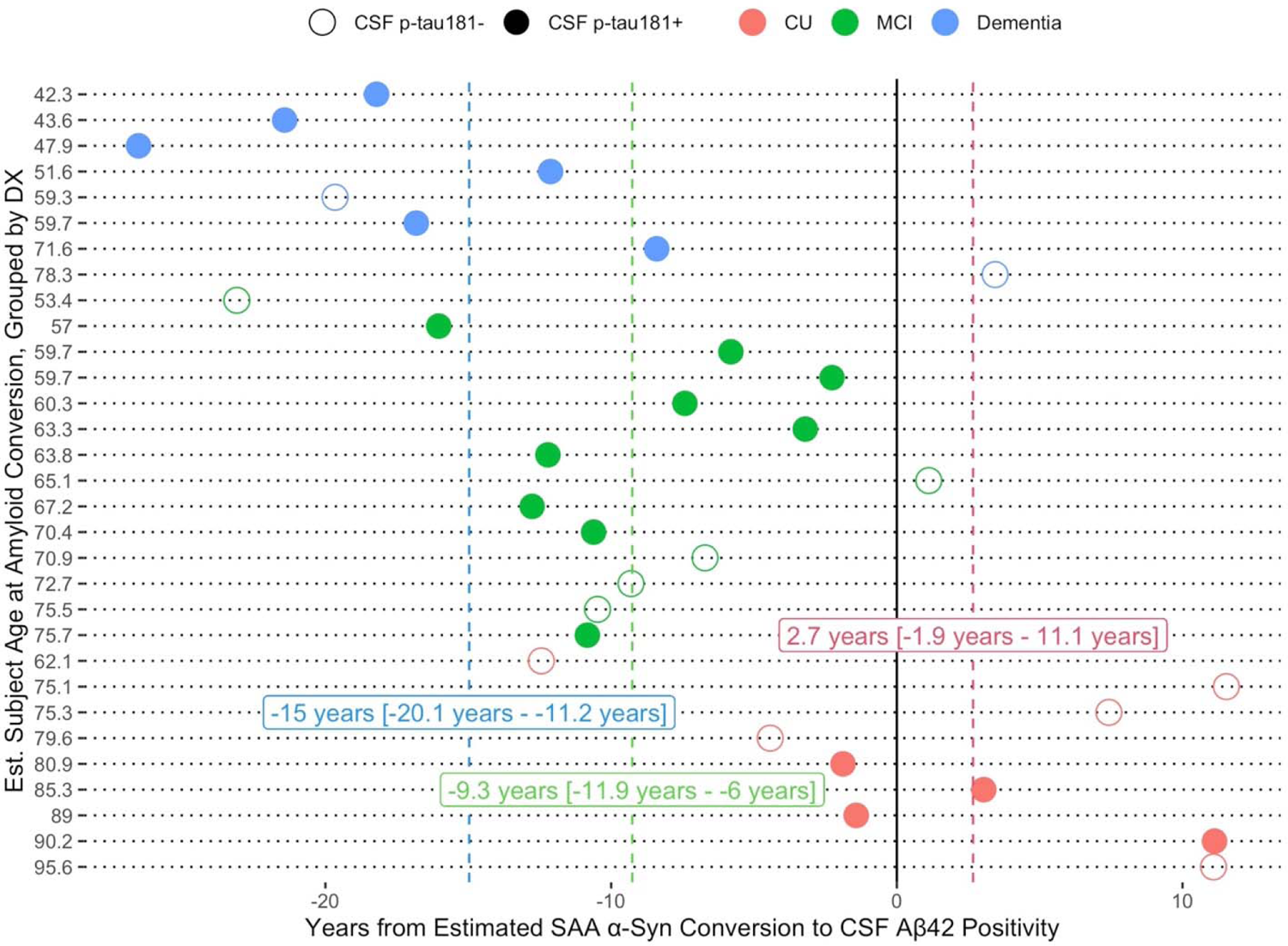
Timing of the CSF Aβ42 pathology (i.e., Aβ) relative to CSF α-syn SAA conversion. Three out of 34 SAA Converters had CSF Aβ42 levels above the upper technical limit of 1700 pg/mL, therefore missing SILA CSF Aβ42 age estimates. Closed and open circles indicate CSF p-tau181 positive and negative participants at the time of SAA conversion, respectively. Vertical dashed lines represent the median (IQR) for timing of the CSF Aβ pathology relative to CSF α-syn SAA conversion time within each diagnostic group.

### Risk for CSF **α**-syn SAA and AD biomarker conversion and change in clinical diagnosis

Risks for CSF α-syn SAA, Aβ42, and p-tau181 biomarker conversion, as well as change in clinical diagnosis (from CU to MCI/Dementia or from MCI to Dementia), were assessed through Cox proportional hazards regression survival analyses.

The survival analysis indicated a significant association of being CSF Aβ42 positive (HR: 2.44; 95% CI: 1.05-5.68) with SAA conversion risk (Figure 5; Supplemental Figure S2). When CSF Aβ42-age and Aβ42-time were used in a repeated survival analysis instead of CSF Aβ42 positivity, the associations remained consistent (data not shown).

**Figure 5:**
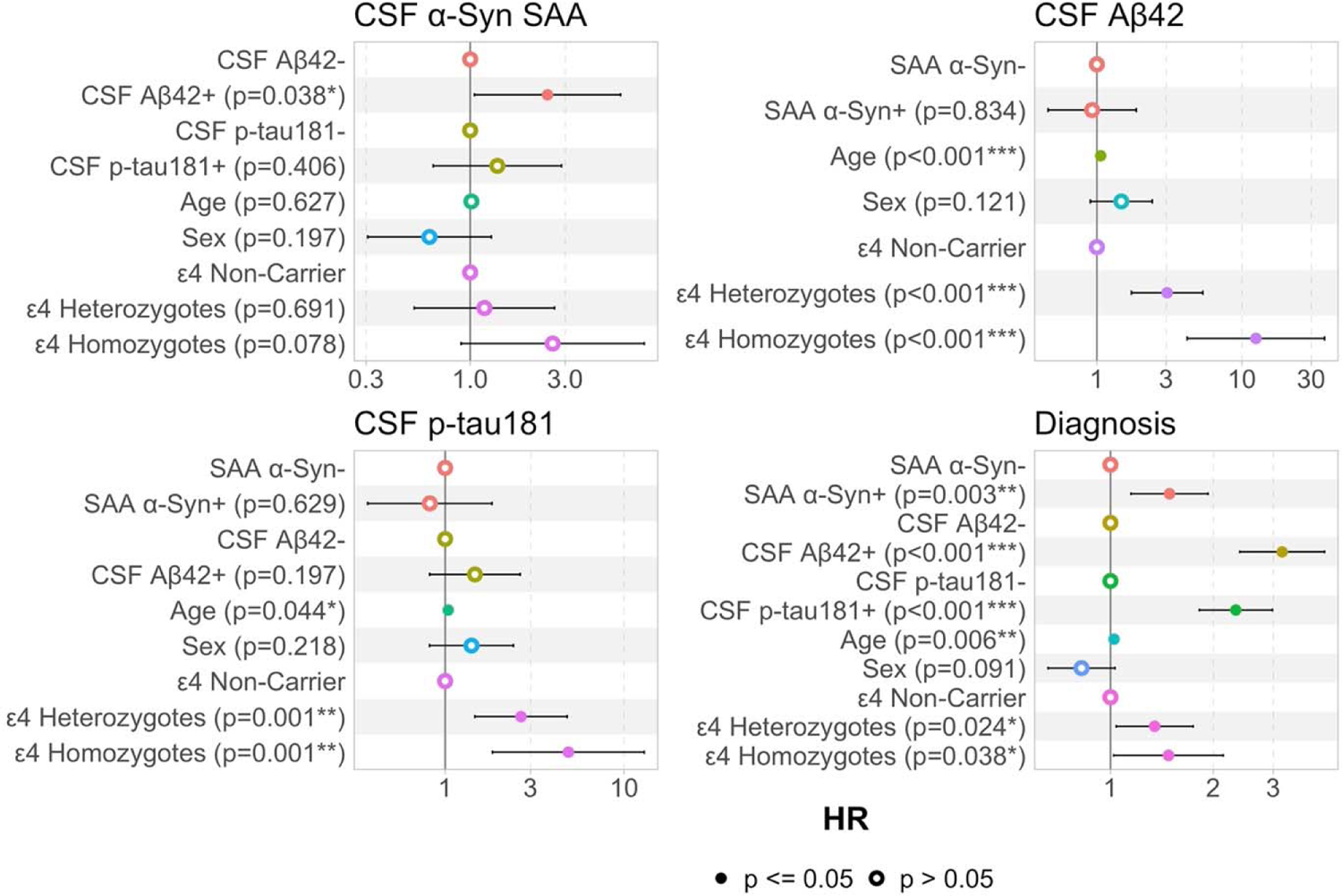
Hazard ratios for predictors from adjusted Cox regression models predicting conversion in CSF α-syn SAA positivity, CSF Aβ42 positivity, CSF p-tau181 positivity, and change in clinical diagnosis (whether from CU to MCI/Dementia or MCI to Dementia).

In contrast, the risk for CSF Aβ42 conversion was associated with older age (HR: 1.06; 95% CI: 1.02-1.10) and *APOE* £4 genotype (Heterozygotes: HR: 3.05; 95% CI: 1.73-5.37; Homozygotes: HR, 12.48; 95% CI, 4.19-37.15) (Figure 5; Supplemental Figure S3). Similarly, the risk for CSF p-tau181 conversion was associated with older age (HR: 1.04; 95% CI, 1.00-1.08), and *APOE* £4 genotype (Heterozygotes: HR, 2.65; 95% CI, 1.46-4.80; Homozygotes: HR, 4.88; 95% CI, 1.84-12.95) (Figure 5; Supplemental Figure S4).

The likelihood of change in clinical diagnosis, whether from CU to MCI/Dementia or MCI to Dementia, was associated with CSF Aβ42 positivity (HR: 3.18; 95% CI: 2.39-4.24), CSF p-tau181 positivity (HR: 2.33; 95% CI: 1.82-2.98), CSF α-syn SAA positivity (HR: 1.49; 95% CI: 1.15-1.93), older age (HR, 1.02; 95% CI, 1.01-1.04), and *APOE* £4 genotype (Heterozygotes: HR: 1.35; 95% CI: 1.04-1.75; Homozygotes: HR, 1.48; 95% CI, 1.02-2.14) as shown in Figure 5 and Supplemental Figure S5.

### Association of SAA kinetic parameters with cohort characteristics and cognitive decline

Within the SAA+ group (N=368), we assessed the independent association of demographic (age, sex, and *APOE* ε4 status) and clinical factors (clinical diagnosis and CSF Aβ42 positivity) with the SAA kinetic parameters (Fmax, Smax, TTT, and TSmax) in a full model including all these factors as well as the total CSF protein concentration. The kinetic parameter analyses were repeated with continuous CSF Aβ42 and p-tau181 levels, cognitive outcome measures, as well as for the associations between change in cognition and change in SAA kinetic parameter, all adjusted for age, sex, *APOE* ε4 status, clinical diagnosis, and total CSF protein concentration,

Although uncorrected association between SAA kinetic parameters and and various demographic (age, sex) and clinical (diagnosis, CSF Aβ42 positivity, CSF p-tau181 levels, ADAS-Cog11, CDR-SB, and language function) factors were observed as illustrated in Figure S6, only a few survived Holm–Bonferroni correction. Specifically, less steep Smax values were associated with older age (β = −0.040; p = 0.0013) and MCI diagnosis compared to being CU or Dementia (β = −0.74; p = 0.0013).

Next, we investigated whether the SAA kinetic parameters were associated with the follow-up time, as an indicator of an association with the duration of α-syn pathology, by utilizing data from individuals who transitioned to SAA positivity from SAA−. Longitudinal kinetic parameters from SAA Converters were aligned at the time of their SAA conversion (i.e., t=0; Figure 6). Following the SAA conversion time, both TTT and TSmax significantly decreased over the subsequent years (p < 0.01), converging to the levels observed within the Stable SAA+ participants. In contrast, Fmax and Smax remained constant over time, at the level observed within the Stable SAA+ participants. This constancy, combined with the decreasing TTT and TSmax, as expected resulted in a significant increase in the AUC-Fluoro (p = 0.03) in years following the SAA conversion time. When repeated within each diagnostic group (i.e., CU, MCI, and Dementia) separately, similar longitudinal profiles were observed (Supplemental Figure S7).

**Figure 6:**
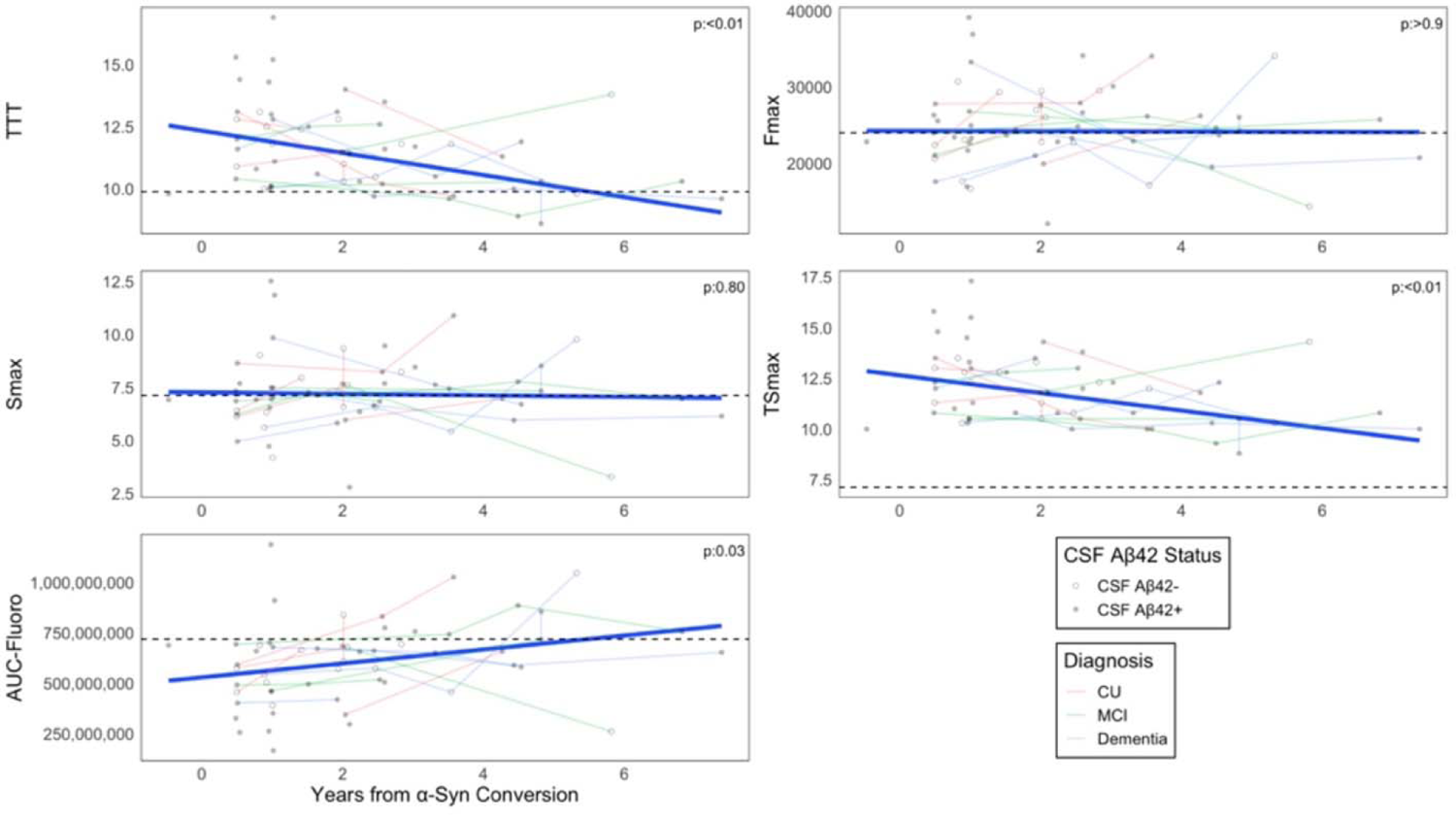
Change in SAA kinetic parameters over years after the SAA conversion time. Horizontal dashed lines represent the average levels of kinetic parameters within Stable SAA+ participants.

## DISCUSSION

We recently applied CSF α-syn SAA to the latest available CSF samples from the ADNI cohort (Phase-1), examining the prevalence of LB pathology (i.e., SAA-positivity) and its correlation with AD biomarkers and cognitive function [19]. Expanding upon this, we incorporated earlier CSF samples (Phase-2) with particular focus on individuals who were SAA+ in Phase-1 (Figure 1). This allowed us to track the progression of CSF Aβ42, α-syn seeds, and p-tau181, along with comprehensive cognitive assessments, in three groups: those with consistent SAA-positivity (Stable SAA+), those with consistent SAA-negativity (Stable SAA−), and those who converted from SAA− to SAA+ status (SAA Converters). The major findings of this study were: 1) SAA+ individuals exhibited a more rapid cognitive decline compared to SAA– individuals, particularly during the MCI stage. SAA+ participants reached a cognitive performance threshold - defined as two standard deviations below the mean of CU individuals - 5.7–9.5 years earlier than their SAA– counterparts. 2) In the subset with longitudinal CSF data, 34 individuals (∼5%) transitioned from SAA– to SAA+ status by their final CSF collection. These ‘SAA Converters’ experienced a more pronounced cognitive decline post-conversion than a matched cohort of Stable SAA− individuals. 3) The risk of converting to SAA+ status was linked to CSF Aβ42-positivity. However, the SAA status itself did not influence the likelihood of becoming positive for either CSF Aβ42 or p-tau181 biomarkers. 4) The positivity in all three CSF biomarkers - Aβ42, p-tau181, and α-syn SAA - independently was associated with greater risk for a change in clinical diagnosis (i.e., CU to MCI/Dementia or MCI to Dementia). Of these, CSF Aβ42 positivity was the strongest risk indicator. 5) The SAA kinetic parameters of Smax was associated with age and MCI diagnosis.

The relationship between SAA status and cognitive trajectories versus change in clinical diagnosis presents a nuanced aspect of AD clinical progression. Most importantly, SAA+ was associated with more rapid cognitive decline in a full adjusted model, predominantly during the MCI stage. We also observed that following SAA conversion, individuals experienced an accelerated decline in cognitive performance. Interestingly, the onset of cognitive impairment, defined as two standard deviations below the mean of CU individuals, for Stable SAA+ compared to Stable SAA– was 5.5–9.4 years earlier for both global and domain-specific cognitive measures. This observation was consistent with previous studies reporting earlier age of symptom onset in AD patients with LB co-pathology [17,36,37]. Our observation that SAA-positivity was a significant risk factor for change in clinical diagnosis, also aligns with findings from these studies. As expected, in the context of AD co-pathologies, CSF Aβ42-positivity as a marker of Aβ-pathology was the strongest risk factor for change in clinical diagnosis.

Our findings provide insights into the interactions particularly of Aβ and α-syn within the AD framework. CSF Aβ42-positivity significantly increased the likelihood of conversion from SAA− to SAA+, supporting the hypothesis that α-syn co-pathology may not arise independently but is rather facilitated by existing Aβ-pathology, especially in symptomatic individuals.

Interestingly, a notable proportion (55%) of CU exhibited SAA-positivity prior to Aβ42-positivity, challenging the linear progression model of AD and suggesting that the temporal sequence and chronicity of AD pathologies may hold greater implications for cognitive decline and clinical diagnosis than the mere presence of multiple pathological entities.

Nevertheless, supporting the hypothesis of Aβ’s influence on α-syn, our findings show that the advent of α-syn pathology appears to be influenced by pre-existing Aβ deposits, with its onset further modulated to some extent by the *APOE* ε4 homozygosity (p=0.078). However, it is important to note that our results do not indicate a significant impact of SAA-positivity on its own in the conversion to biomarker positivity for CSF Aβ42 or p-tau181. This suggests that the effect of Aβ on α-syn is likely unidirectional, without evidence of a reciprocal relationship.

Consistent with neuropathological evidence from autopsy studies [2,5,6,38], which show a higher prevalence of α-syn changes in brains with abundant neuritic plaques but not necessarily correlating with the severity of neurofibrillary tangles, our findings did not identify a strong association between SAA conversion and CSF p-tau181-positivity, as a tau biomarker. We previously reported an inverse relationship between SAA+ prevalence and CSF p-tau181 levels and flortaucipir PET burden in the Dementia stage of the disease [19]. Consistently, although it did not reach significance due to limited sample size (p=0.12; Supplemental Figure S8), the risk for SAA conversion within CSF Aβ42+ Dementia was marginally associated with lower levels of CSF p-tau181 (HR: 0.21; 95% CI: 0.03-1.50). Immunostaining of brain tissues for tau and α-syn antibodies has revealed a higher burden of α-syn pathology in AD with LBs (AD+LB) compared to PD dementia cases. Interestingly, the pathological tau load was found to be similar or even slightly lower in AD+LB compared to AD alone, with co-localization of phosphorylated tau and α-syn within astrocytes in the middle temporal gyrus [39]. In contrast, autopsy literature on PD frequently reports concurrent deposition of α-syn and tau, reflecting a complex interplay that may differ from AD pathology. Reviews such as that by Twohig and Nielsen [37] posit that α-syn may interact more significantly with tau than with Aβ, emphasizing the importance of protein species, whether soluble or insoluble, in the early seeding events of these pathologies. This suggests a potential divergence in the pathophysiological mechanisms underlying these proteinopathies in different neurodegenerative disease presentations.

Taken together, our findings suggest possible interactions between AD and LB pathologies, potentially involving crosstalk mechanisms and genetic predispositions such as the *APOE* ε4 allele. These interactions may be influenced by compromised proteostasis, raising the question of whether α-syn preferentially engages with one pathological species over another, or if it is merely the timing of the emergence of these pathologies that dictates their interrelationship. It is important to note that our study was not designed or powered to understand the mechanism by which Aβ and α-syn as well as tau pathologies interact and coexist in the brain of AD cases.

Nevertheless, prior work has suggested that interaction between Aβ and α-syn may reduce protein clearance, activate inflammatory processes, increase tau phosphorylation, and enhance aggregation of each other [40]. Notably, the biomarkers currently utilized for detecting these pathological changes may have inherent limitations, such that measurable changes in CSF Aβ42 levels may precede alterations in p-tau levels.

Our findings revealed that following SAA conversion, the kinetic parameters TTT and TSmax significantly decreased over subsequent years, converging to levels observed within the Stable SAA+ participants. However, the Fmax and Smax remained constant over time, at levels similar to those observed within the Stable SAA+ participants. One interpretation of our findings could be a potential association between amplification time at the emergence of LB pathology in the context of co-pathologies while in later stages SAA features remain stable over time. The faster aggregation kinetics with time from the initial SAA-positivity might also be related to many factors including: changing number of α-syn aggregates over time, changes in the biophysical properties of the seeds, as well as presence of lipids, proteins or other compounds in the CSF. The qualitative nature of the current SAA protocol though poses a limitation in assigning SAA-positivity time to Stable SAA+ cases, potentially introducing an unknown source of heterogeneity to the kinetic parameter measures. Nevertheless, within the Stable SAA+ group, Fmax and Smax were associated with age, sex, CSF Aβ42 positivity, MCI diagnosis, and CSF p-tau181 levels. Future longitudinal studies with larger sample sizes are warranted to further understand whether the associations between SAA kinetic parameters with these demographic and AD characteristics are reflections of a widespread α-syn-driven LB pathology and a greater leakage of misfolded α-syn species from degenerating neurons. Accordingly, recent studies suggest that SAA kinetic parameters, particularly TTT and TSmax, were associated with clinical and cognitive characteristics of PD and DLB patients, measured by Unified Parkinson’s disease rating scale part (UPDRS) III and Montreal Cognitive Assessment [41,42]. Although not survived correction for multiple comparison, TSmax was associated with cognitive impairment (ADAS-Cog11, CDR-SB, and language function) and decline (language domain). TSmax might be inversely proportional to the density of α-syn seeds, therefore a faster amplification of α-syn seeds might again be a reflection greater leakage of misfolded α-syn species from degenerating neurons underlying greater cognitive worsening. Another crucial reason that warrants further investigation is the extent to which LB-pathology contributes to clinical and cognitive impairment (i.e., quantitative burden of LB-pathology) in a dose-specific manner might be influenced by its primary pathology status versus co-pathology status.

Limitations of the study include the exclusion of individuals with prominent DLB clinical features from ADNI, lack of measures associated with PD/DLB clinical features (e.g. UPDRS and smell tests), as well as the limited ancestral diversity in the available ADNI cohorts, and SAA kinetic parameters not quantified for SAA– participants. In addition, Aβ-PET and tau-PET imaging, gold-standard biomarkers for AD Aβ and tau pathologies, were only available for a limited number of study participants (51% and 28%, respectively).

## CONCLUSION

In summary, our results highlight the potential for interplay between Aβ and α-syn and their impact on disease progression, emphasizing the importance of further investigation into their underlying mechanisms in the context of co-pathologies of AD. The longitudinal tracking of SAA+ alongside other biomarkers prompts consideration of differential diagnosis between AD and other neurodegenerative conditions, especially DLB. Moving forward, it is imperative to broaden the detection of LB-pathology in diverse cohorts to enhance our understanding of the causes and triggers of AD and LB co-pathologies.

## Data Availability

The ADNI data used in this study were obtained from the ADNI database (https://adni.loni.usc.edu). All ADNI data are shared without embargo through the LONI Image and Data Archive (IDA).

## DATA SHARING

The ADNI data used in this study were obtained from the ADNI database (https://adni.loni.usc.edu). All ADNI data are shared without embargo through the <u>LONI Image</u> <u>and Data Archive</u> (IDA).

## ACKNOWLEDGEMENTS

This work was supported in part by the Intramural Research Program of the National Institute on Aging (NIA), and the Center for Alzheimer’s and Related Dementias (CARD), within the Intramural Research Program of the National Institute on Aging and the National Institute of Neurological Disorders and Stroke (AG000546). Data collection and sharing for this project was funded by the Alzheimer’s Disease Neuroimaging Initiative (ADNI) (National Institutes of Health Grant U01 AG024904). ADNI is funded by the National Institute on Aging, the National Institute of Biomedical Imaging and Bioengineering, and through generous contributions from the following: AbbVie, Alzheimer’s Association; Alzheimer’s Drug Discovery Foundation; Araclon Biotech; BioClinica, Inc.; Biogen; Bristol-Myers Squibb Company; CereSpir, Inc.; Cogstate; Eisai Inc.; Elan Pharmaceuticals, Inc.; Eli Lilly and Company; EuroImmun; F. Hoffmann-La Roche Ltd and its affiliated company Genentech, Inc.; Fujirebio; GE Healthcare; IXICO Ltd.; Janssen Alzheimer Immunotherapy Research & Development, LLC.; Johnson & Johnson Pharmaceutical Research & Development LLC.; Lumosity; Lundbeck; Merck & Co., Inc.; Meso Scale Diagnostics, LLC.; NeuroRx Research; Neurotrack Technologies; Novartis Pharmaceuticals Corporation; Pfizer Inc.; Piramal Imaging; Servier; Takeda Pharmaceutical Company; and Transition Therapeutics. The Canadian Institutes of Health Research is providing funds to support ADNI clinical sites in Canada. Private sector contributions are facilitated by the Foundation for the National Institutes of Health (www.fnih.org). The grantee organization is the Northern California Institute for Research and Education, and the study is coordinated by the Alzheimer’s Therapeutic Research Institute at the University of Southern California. ADNI data are disseminated by the Laboratory for Neuro Imaging at the University of Southern California.

## CONSENT STATEMENT

ADNI study was approved by each ADNI study site’s respective institutional review board and informed written consents were obtained from all participants.

