## Supplemental Material for "Association of CSF α-Synuclein Seed Amplification Assay Positivity with Disease Progression and Cognitive Decline: A Longitudinal Alzheimer’s Disease Neuroimaging Initiative Study"

**SUPPLEMENTAL MATERIALS**

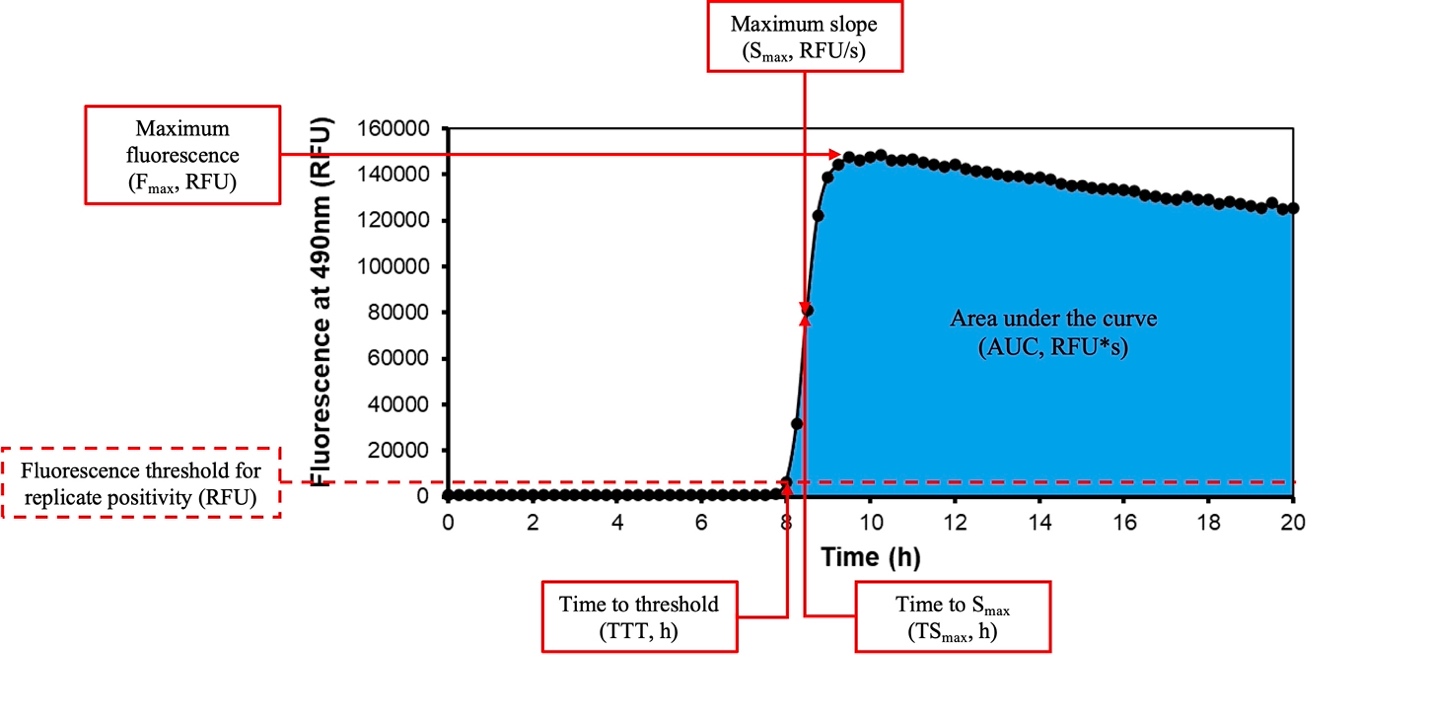

**Figure S1**. Illustration of the five kinetic parameters estimated for each positive replicates: 1) Time to Threshold (TTT, [hours]) – time in hours when the fluorescence signal reaches the lower patient classification threshold (1000 RFU); 2) Maximum Fluorescence (Fmax, [RFU]) – maximum of the reaction signal in relative fluorescence units (RFU); 3) AUC-Fluoro (RFU, [seconds]) – area under the signal versus time reaction curve in RFU; 4) Maximum Slope (Smax, [RFU, seconds]) – maximum of the derivative of the signal/time reaction curve in RFU/seconds; 5) Time to Smax (TSmax, [hours]) – the time in hours when the maximum slope occurs.

| **Table S1:** Missing data counts and percentages for clinical and biomarker data within study groups. | | | | |
| --- | --- | --- | --- | --- |
| Characteristic | SAA- Stable, N = 641*^1^* | SAA+ Stable, N = 150*^1^* | Converters (Before), N = 34*^1^* | Converters  (After), N = 34*^1^* |
| CSF Aβ42 | 36 (5.8%) | 0 (0%) | 1 (2.9%) | 3 (2.0%) |
| CSF p-tau181 | 36 (5.8%) | 0 (0%) | 1 (2.9%) | 3 (2.0%) |
| ADAS13 | 36 (5.8%) | 0 (0%) | 1 (2.9%) | 3 (2.0%) |
| CDRSB | 14 (2.3%) | 1 (2.9%) | 1 (2.9%) | 3 (2.0%) |
| PACC | 11 (1.8%) | 0 (0%) | 0 (0%) | 2 (1.3%) |
| MMSE | 5 (0.8%) | 0 (0%) | 1 (2.9%) | 0 (0%) |
| Memory composite | 8 (1.3%) | 0 (0%) | 1 (2.9%) | 0 (0%) |
| Executive function | 8 (1.3%) | 0 (0%) | 1 (2.9%) | 1 (0.7%) |
| Language composite | 6 (1.0%) | 0 (0%) | 0 (0%) | 0 (0%) |
| *^1^* n (%) | | | | |

| **Table S2:** Clinical diagnosis transitions among SAA Converters | | | |
| --- | --- | --- | --- |
|  | Before Conversion, N = 34*^1^* | After Conversion, N = 34*^1^* | Most Recent, N = 34*^1^* |
| CU | 11 (32%) | 11 (32%) | 12 (35%) |
| MCI | 15 (44%) | 10 (29%) | 6 (18%) |
| Dementia | 8 (24%) | 13 (38%) | 16 (47%) |
| *^1^* n (%) | | | |

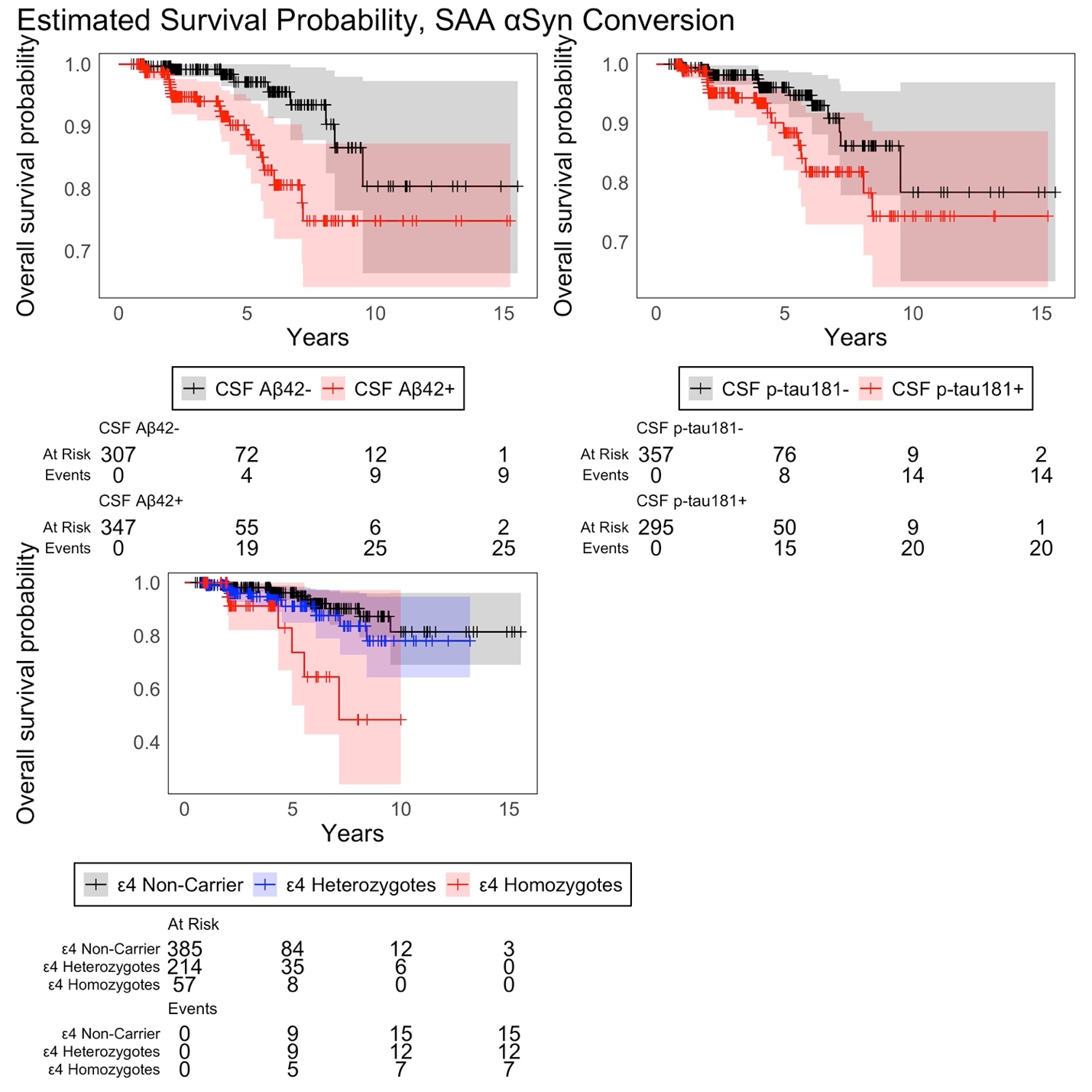

**Figure S2**: Kaplan-Meier estimates of survival for CSF α-syn SAA phenotype change, stratified by CSF Aꞵ42 positivity, CSF p-tau181 positivity, and *APOE* ε4 genotype.

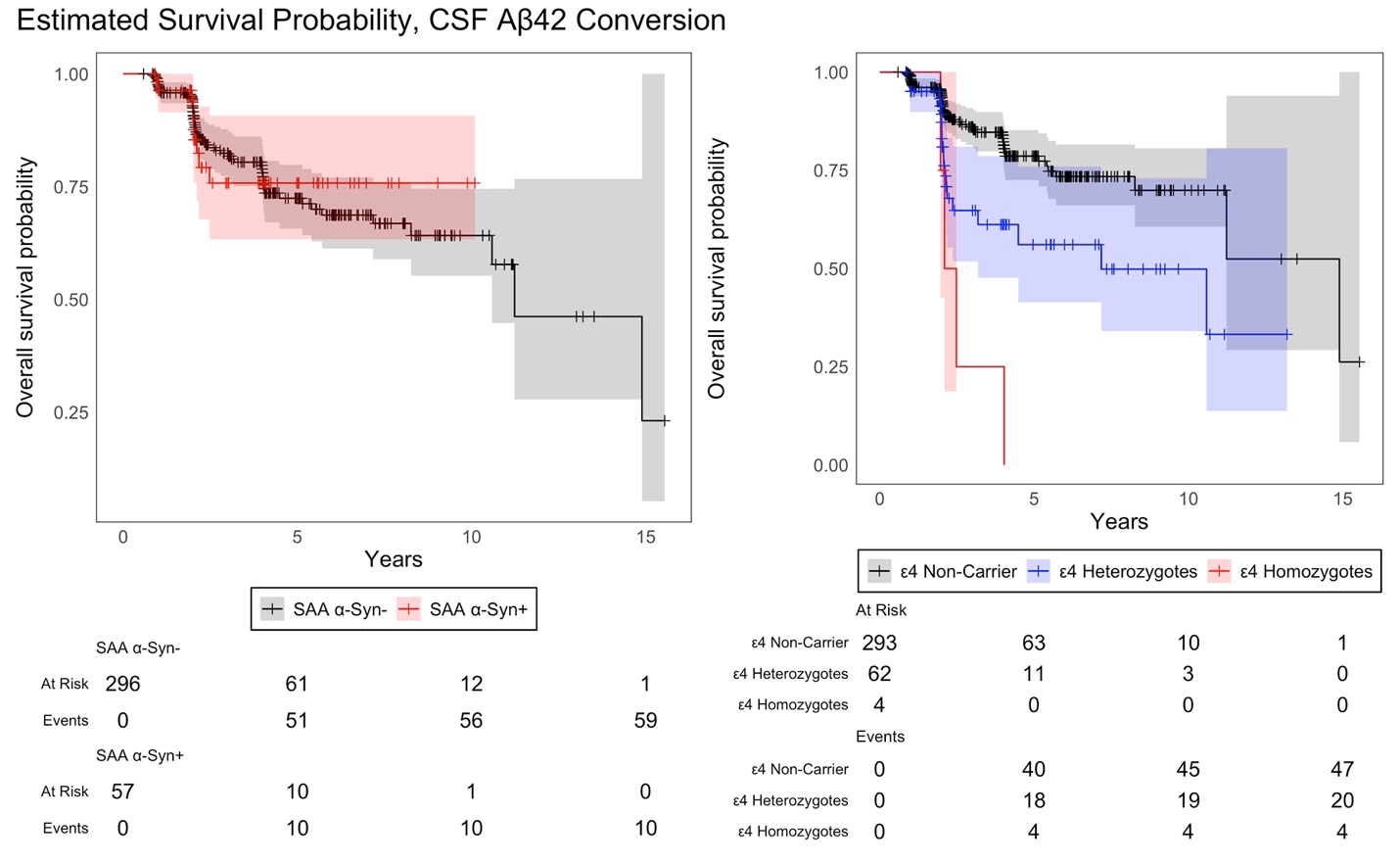

**Figure S3**: Kaplan-Meier estimates of survival for CSF Aꞵ42 phenotype change, stratified by

CSF α-syn SAA positivity, and *APOE* ε4 genotype.

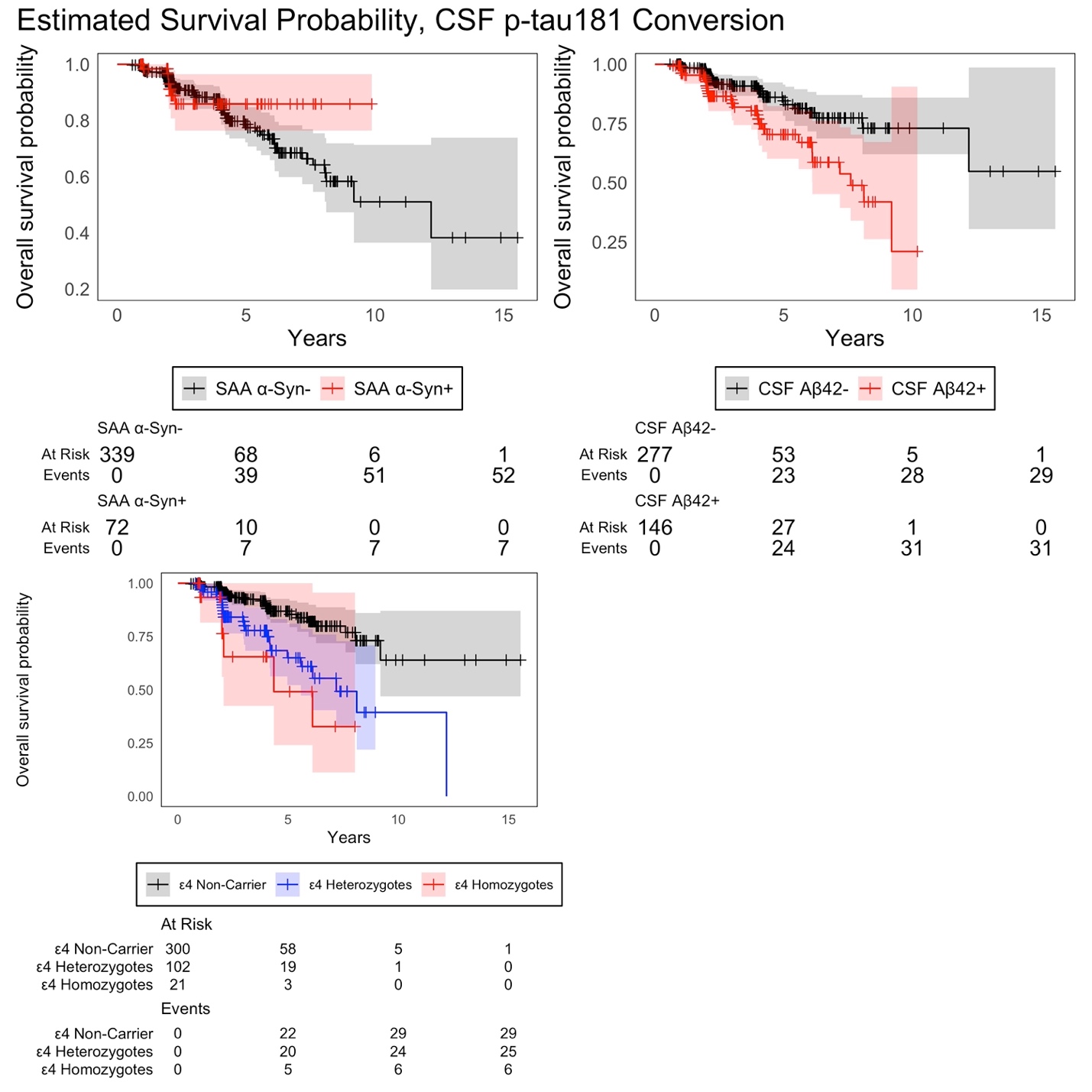

**Figure S4**: Kaplan-Meier estimates of survival for CSF p-tau181 phenotype change, stratified by CSF α-syn SAA positivity, CSF Aꞵ42 positivity, and *APOE* ε4 genotype.

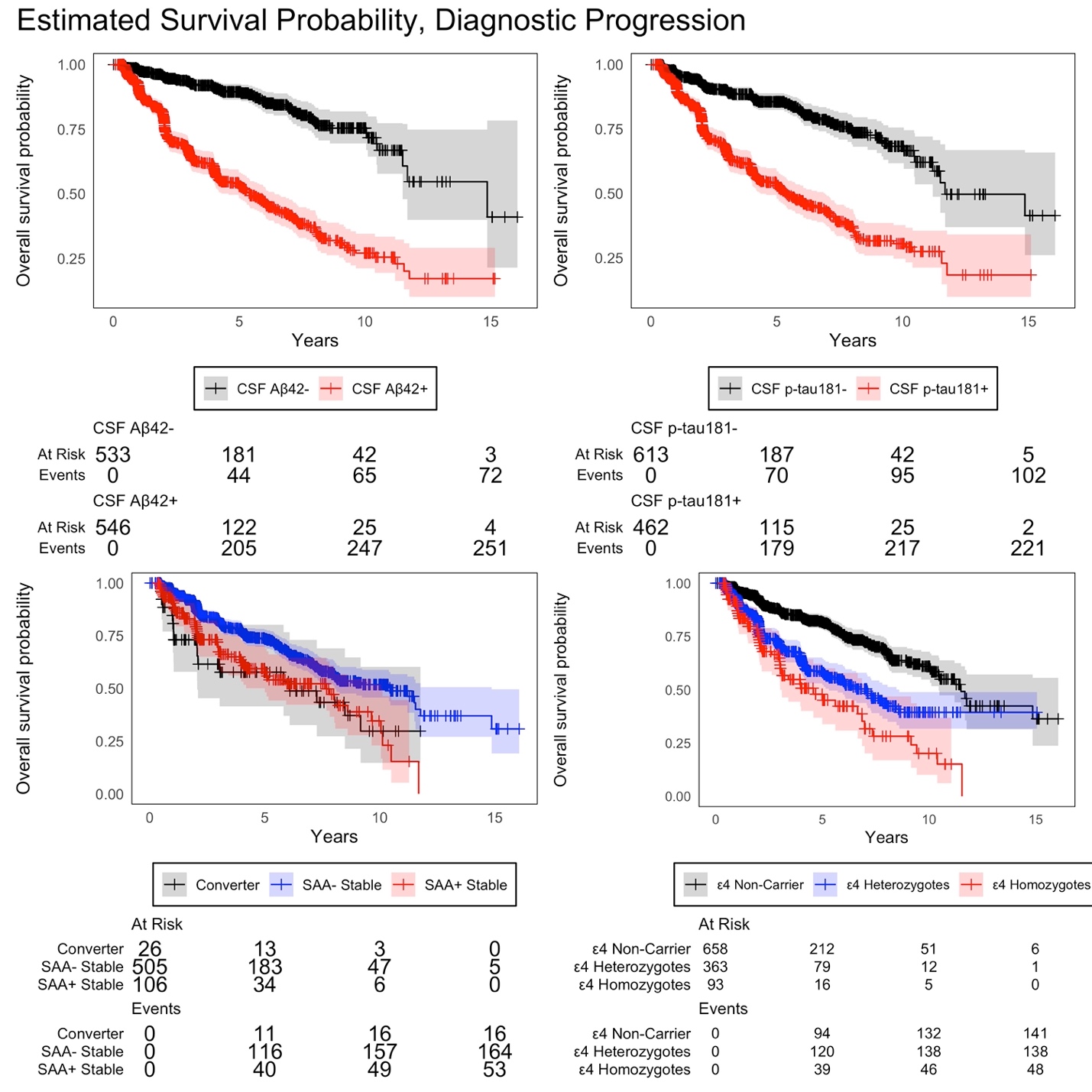

**Figure S5**: Kaplan-Meier estimates of survival for clinical progression to MCI and Dementia diagnosis, stratified by CSF Aꞵ42 positivity, CSF p-tau181 positivity, CSF α-syn SAA positivity, and *APOE* ε4 genotype.

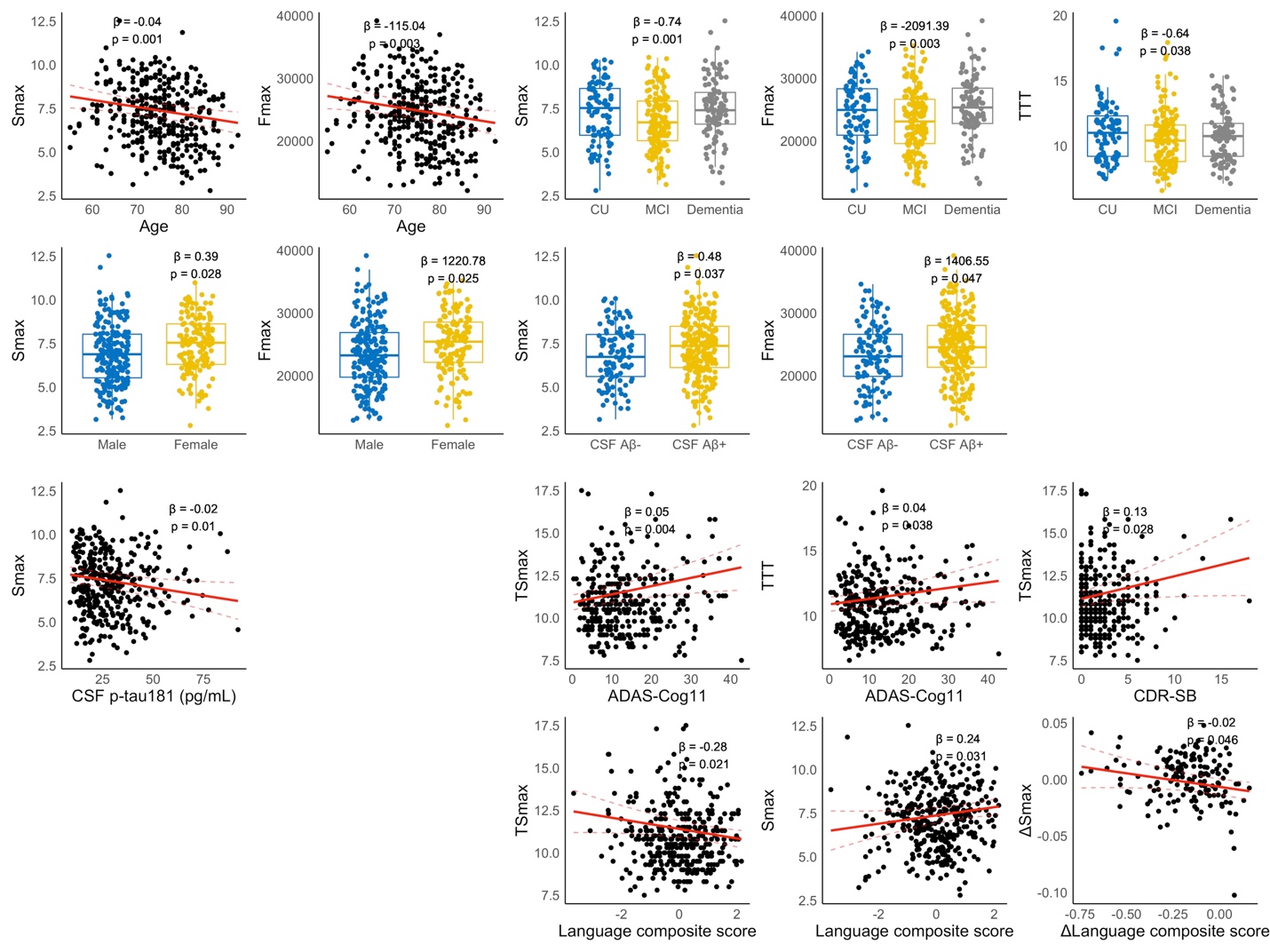

**Figure S6:** SAA kinetic parameters associated with cohort characteristics and disease progression.

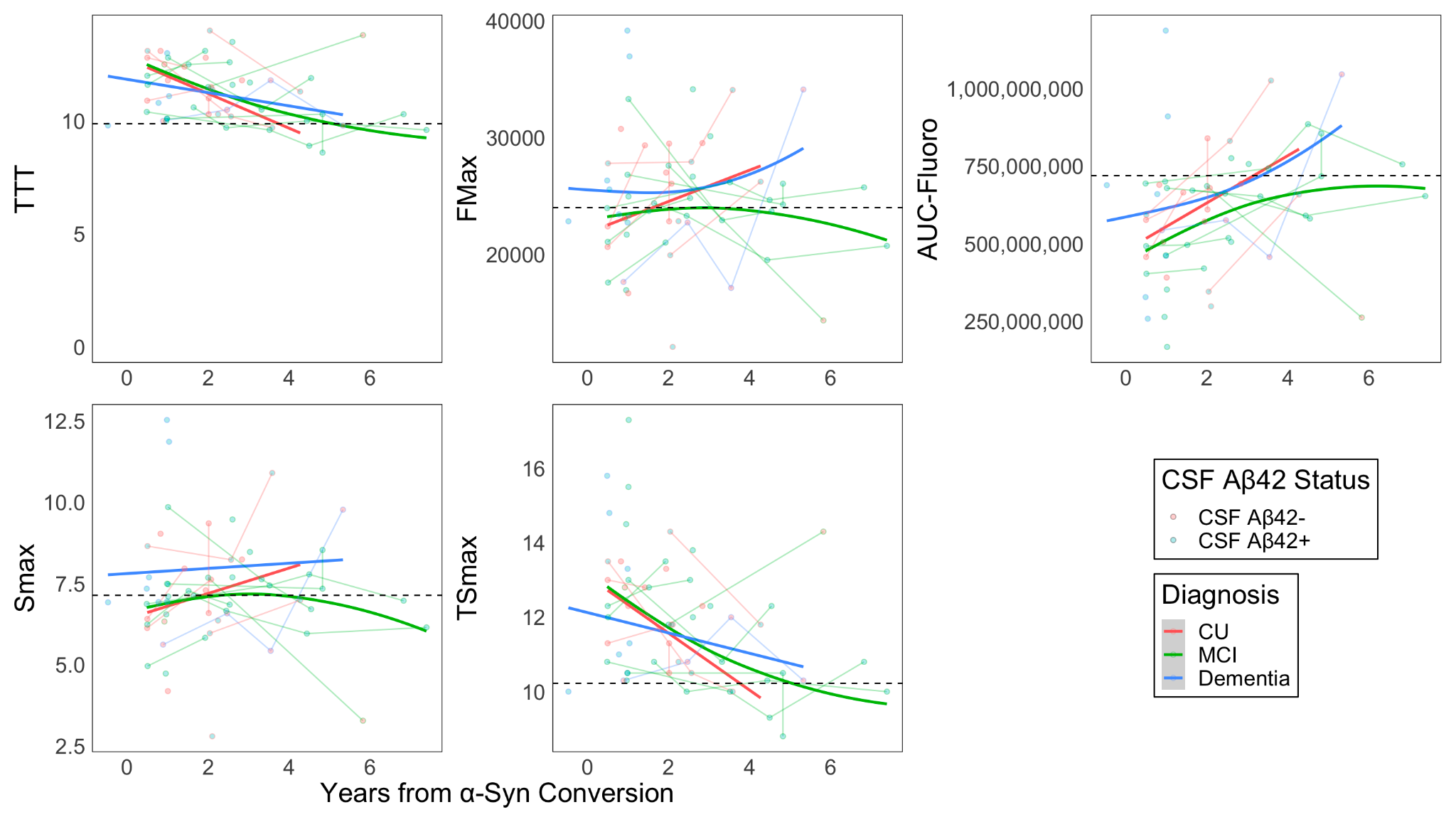

**Figure S7:** GAM-estimated cross-sectional associations of years from estimated α-syn SAA conversion with kinetic parameters by diagnosis and CSF Aβ42 status.

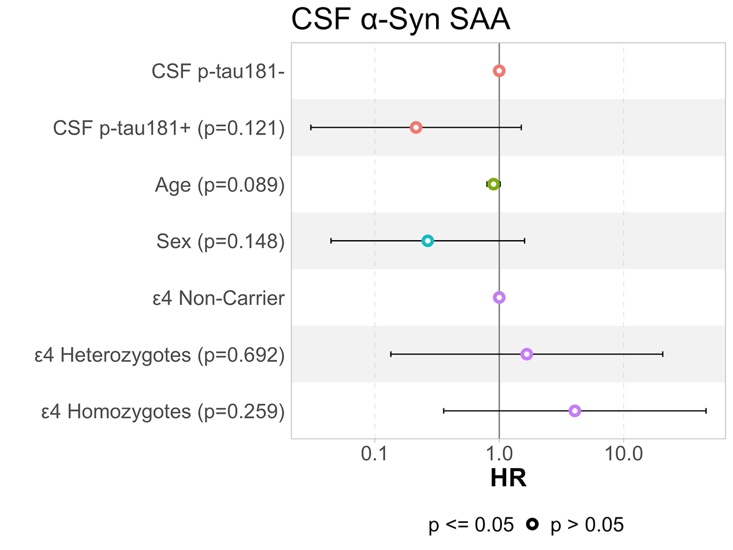

**Figure S8:** Hazard ratios for predictors from adjusted Cox regression models predicting conversion in CSF α-syn SAA positivity within individuals with CSF Aβ42+ Dementia.
